# High-altitude exercise orchestrates a divergent immune landscape: cytotoxic suppression, humoral compensation, and neutrophil functional reprogramming

**DOI:** 10.64898/2026.09.17.26363363

**Authors:** Junlei Zhang, Jiaqi Wang, Ruoyi Xue, Yutong Dong, Zujie Tang, Chen Zhang, Wubin Yang, Yangkai Zhang, Guangxinghao Zhang, Manying Guo, Rui Jian, Yi Huang, Yanping Tian, Yan Ruan, Yan Hu

**Author notes:** These authors contributed equally to this work (Junlei Zhang, Jiaqi Wang, Ruoyi Xue). Corresponding author. (Y.T); (Y.R); (Y.H).

## Abstract

Physical exertion at high-altitude imposes a dual stress of hypoxia and mechanical load, yet the underlying immune adaptations remain poorly understood. Plasma proteomics and single-cell RNA sequencing were performed in 46 healthy men undergoing standardized exercise at low and high altitudes, and the findings were validated in mice exposed to chronic extreme hypoxia (simulated 5,800 m) combined with exhaustive exercise. Whereas broad immune activation was induced by low-altitude exercise, cytotoxic and innate effector programs in CD8 T cells, natural killer cells, monocytes, and dendritic cells were broadly suppressed following high-altitude exercise. In contrast, humoral immunity was enhanced, as indicated by the activation of plasmablasts and CD4 memory and proliferating T cells. Mature neutrophils were functionally reprogrammed toward phagocytosis and degranulation, accompanied by an expansion of activated circulating neutrophils. A high-altitude-specific program involving chemotaxis and vascular extravasation was further identified by plasma proteomic and murine muscle transcriptomic analyses. These adaptations were associated with enhanced APRIL, MIF, and Annexin A1 signaling and attenuated CCL-chemokine and complement pathways. Collectively, high-altitude exercise was shown to induce coordinated immune reconfiguration characterized by cellular immunosuppression, selective humoral enhancement, and neutrophil-mediated tissue infiltration.

## Introduction

Globally, approximately 81.6 million people live at or above 2,500 m (1,54), and many residents, workers, and travelers perform physically demanding activity in these environments^1^. Physical exercise, which is a potent physiological stressor, elicits immediate and transient changes in metabolic demands, hormone levels, as well as immune cell redistribution and function^2–5^. When strenuous exercise is superimposed on chronic high-altitude exposure, hypoxia and exertion may interact to alter cardiovascular, metabolic, and immune homeostasis and may further reduce exercise capacity^6, 7^. Exploring the underlying mechanisms by which exercise stress following high-altitude acclimatization impacts systemic homeostasis is crucial not only for safeguarding health and performance among high-altitude populations, but also for understanding the limits of human adaptation and the regulatory networks underlying combined extreme stresses^8–10^.

At low altitude, acute exercise causes rapid redistribution of circulating leukocyte subsets, with preferential mobilization of highly differentiated and cytotoxic NK- and CD8 T-cell populations, together with transient changes in the circulating concentrations of cytokines and other soluble mediators^11^. These responses reflect coordinated neuroendocrine, hemodynamic, and immune regulation; their functional consequences depend on exercise intensity, duration, and recovery. Our previous research has shown that chronic high-altitude exposure induces a “remodeling pattern” in the immune system characterized by innate immune activation and adaptive immune suppression. This pattern includes accelerated neutrophil maturation and enhanced cytotoxicity of CD56^dim^ NK cells, accompanied by suppressed plasmablast maturation and diminished T cell immune responsiveness. Physical exercise performed in high-altitude or hypoxic environments imposes a synergistic physiological stress from both exertion and oxygen deprivation, which may lead to more pronounced immunological alterations. Recently, a multi-omics study has revealed that during extreme high-altitude mountaineering, the immune system exhibits activated inflammatory responses and impaired immune effector functions, alongside an enhanced cellular response to hypoxia and oxidative stress^12^. However, the immune system’s response to acute exercise stress following long-term high-altitude acclimatization, and its underlying cellular and molecular regulatory mechanisms, are still poorly understood.

To address these limitations, we designed a study in which participants underwent an identical, standardized maximal exercise test both at sea level and after long-term acclimatization at 3650 meters^13^. Blood samples collected at pre-exercise and within 15 minutes post-exercise were analyzed using single-cell transcriptomics and quantitative plasma proteomics. To robustly validate these human multi-omics findings, we further established a murine model subjected to chronic extreme hypoxia (simulated 5800 m) and exhaustive exercise. By integrating human discovery with murine phenotypic validation, we aim to map the immune system’s complex adaptive landscape under the combined stress of chronic hypoxia and acute exercise, providing crucial insights for mitigating altitude-induced immune dysfunction and optimizing acclimatization strategies.

## Results

### 1. Physiological parameters and Immune Profiling of the subjects in response to low- and high-altitude exercise

Participants underwent the same standardized 12-min exercise protocol at low altitude and after 90 days of acclimatization to 3,650 m, with peripheral blood collected before and within 15 min after exercise (Figure 1A). Physiological and hematological measurements showed expected adaptations to chronic hypoxia, including reduced resting oxygen saturation, increased heart rate, and enhanced erythropoiesis (Supplementary Tables 1-2). Acute exercise increased circulating leukocytes at both altitudes, including neutrophils, lymphocytes, and monocytes. Several altitude-dependent differences were also observed: red blood cell count and hemoglobin decreased slightly after exercise at high altitude but remained relatively stable at low altitude, whereas baseline creatine kinase levels were higher after high-altitude acclimatization.

**Figure 1.**
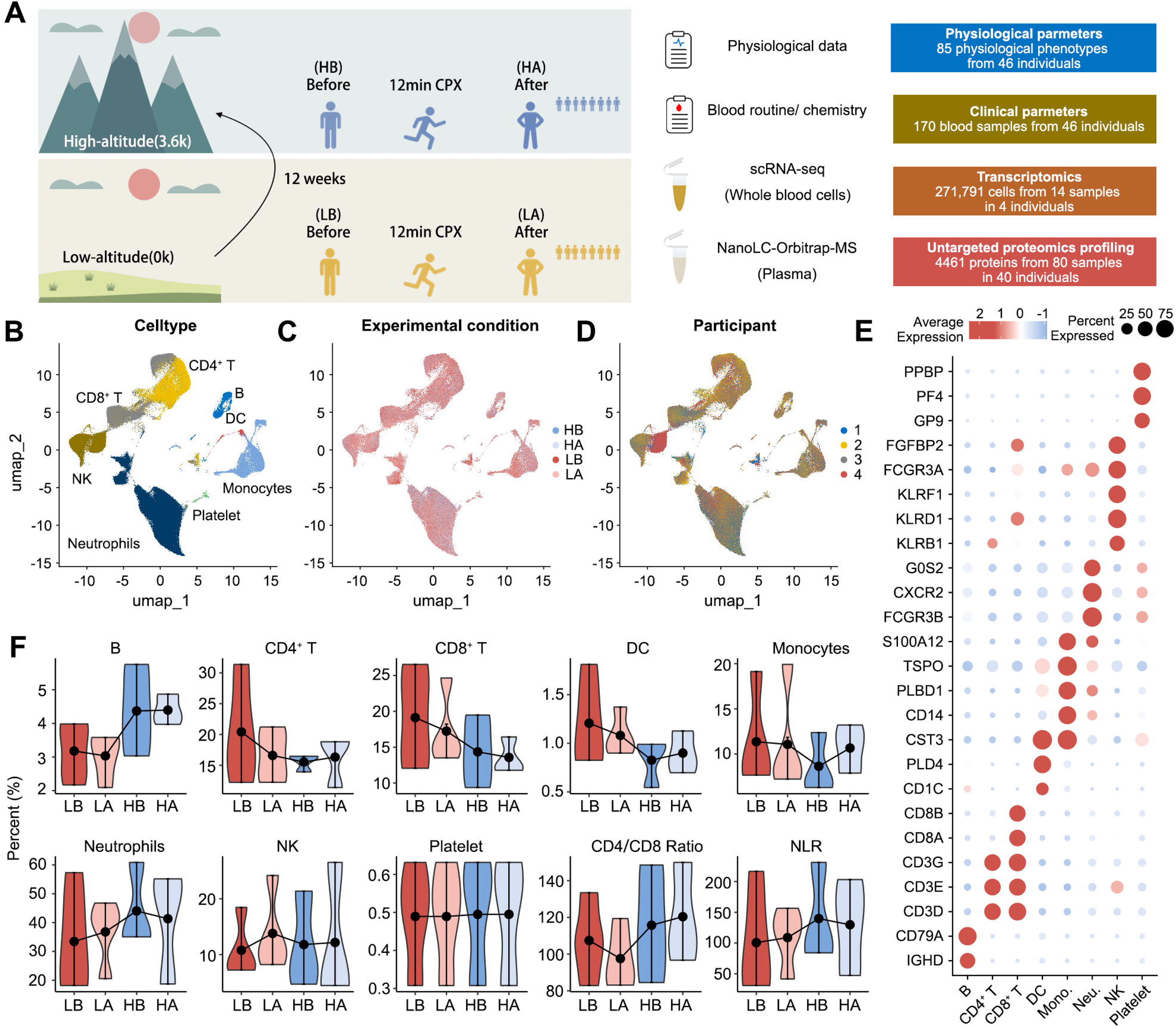
Comprehensive immune-proteomics profiling of exercise at low-altitude and high-altitude. A. Schematic overview of the experimental design and sample collection. B. UMAP analysis showing integrated patterns of eight immune cell types. C. UMAP analysis showing integrated patterns of four experimental conditions. D. UMAP analysis showing integrated patterns of sample attributes. E. Dot plots showing the percentages and average expressions of marker genes across eight immune cell types. F. Violin plots showing the changes in the relative proportions of immune cell types.

Single-cell RNA sequencing generated 271,791 transcriptomes covering 20,482 genes. Eight major cell populations were identified, including B cells, CD4^+^ T cells, CD8^+^ T cells, dendritic cells (DCs), monocytes, NK cells, neutrophils, and platelets (Figure 1B-E). Acute exercise induced rapid immune-cell redistribution at both altitudes (Figure 1F). At low altitude, the proportions of CD4^+^ T cells, CD8^+^ T cells, monocytes, and DCs decreased, whereas neutrophils and NK cells increased. In contrast, several responses after high-altitude exercise, including changes in CD4^+^ T cells, monocytes, DCs, neutrophils, the CD4^+^/CD8^+^ T-cell ratio, and the neutrophil-to-lymphocyte ratio, occurred in the opposite direction. Thus, chronic high-altitude exposure markedly altered the pattern of exercise-induced immune-cell redistribution.

### 2. Divergent transcriptional landscapes in response to low- and high-altitude exercise

To define the molecular responses to exercise at different altitudes, transcriptional changes were compared across major immune-cell populations (Figure 2A-C; Supplementary Table 3). Low-altitude exercise predominantly induced gene upregulation across immune lineages, consistent with broad immune activation. In contrast, high-altitude exercise was characterized by widespread transcriptional downregulation, with neutrophils representing a notable exception.

**Figure 2.**
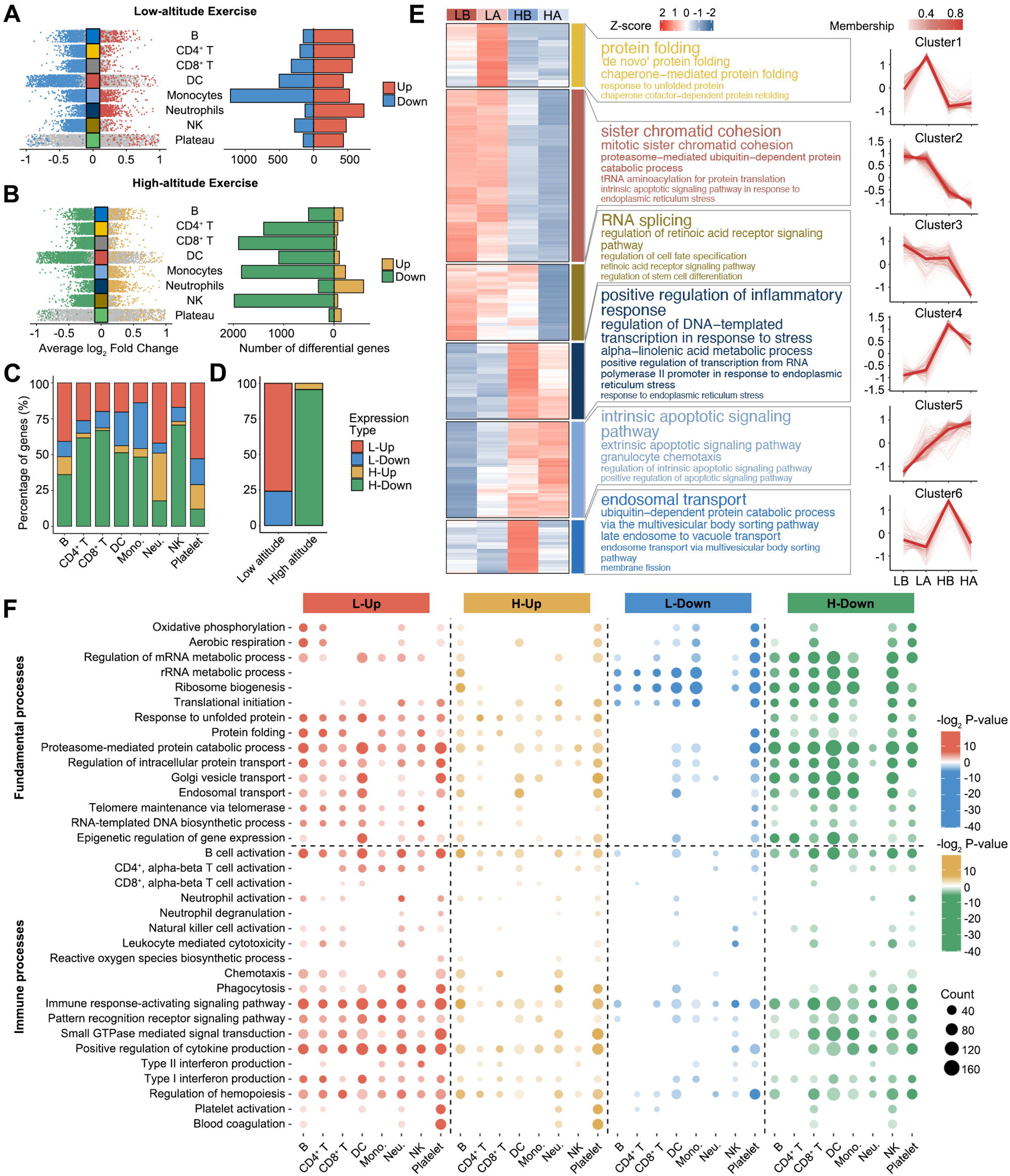
Single-cell RNA sequencing reveals exercise-induced immune landscape remodeling at low-altitude and high-altitude. A. Multi-group volcano plots displaying DEGs in eight immune cell types induced by exercise at low-altitude, with significantly upregulated (red) and downregulated (blue) genes indicated. B. Multi-group volcano plots displaying DEGs in eight immune cell types induced by exercise at high-altitude, with significantly upregulated (orange) and downregulated (green) genes indicated. C. Stacked bar plot showing the relative proportion of four types of DEGs in each immune cell type: low-altitude upregulated (L-Up, red), low-altitude downregulated (L-Down, blue), high-altitude upregulated (H-Up, orange), and high-altitude downregulated (H-Down, green). D. Bar plot comparing the percentage distribution of common DEGs that were consistently regulated across multiple immune cell types at low-altitude and high-altitude. E. Mfuzz clustering analysis of common DEGs based on expression trajectories across four experimental conditions (LB, LA, HB, HA), revealing six distinct temporal patterns (C1-C6). Left panel shows the heatmap of gene expression Z-scores; middle panel displays GO enrichment analysis for each cluster; right panel illustrates the membership-weighted expression trends for each cluster. F. Functional enrichment analysis of DEGs across eight immune cell types under low-altitude and high-altitude exercise conditions. The bubble plot displays significantly enriched GO biological processes for four categories: L-Up (red), H-Up (orange), L-Down (blue), and H-Down (green). Color intensity represents -log_2_(P-value), and bubble size indicates the number of genes enriched in each GO term.

Analysis of shared exercise-responsive genes further demonstrated distinct expression trajectories across the four experimental conditions (Figure 2D-E). Several cellular maintenance programs, including cell-cycle regulation, protein metabolism, RNA processing, and protein-folding pathways, were progressively suppressed after high-altitude exposure and exercise. In contrast, pathways associated with apoptosis and granulocyte chemotaxis showed progressive activation and reached their highest levels after high-altitude exercise.

Functional enrichment analysis revealed a similarly divergent pattern across immune lineages (Figure 2F). Low-altitude exercise broadly enhanced antiviral responses, cytokine production, and other immune and cellular-maintenance processes. High-altitude exercise, however, suppressed pattern-recognition, cytokine-production, and cytotoxicity programs in CD8^+^ T cells, NK cells, and monocytes. B cells and CD4^+^ T cells retained selected activation-associated programs, whereas neutrophils showed a distinct functional profile characterized by enhanced chemotaxis, degranulation, reactive oxygen species metabolism, and translational activity despite suppression of cytokine- and interferon-related signaling. Together, these findings identify a high-altitude-specific immune response characterized by attenuation of cytotoxic and innate effector programs, selective preservation of humoral-associated activity, and neutrophil functional reprogramming.

### 3. High-altitude exercise selectively activates plasmablasts and CD4^+^ memory/proliferating T cells to enhance humoral immunity

#### 3.1 Effects of Low- and High-Altitude Exercise on B-Cell Differentiation and Function

Four B-cell subsets were identified on the basis of canonical markers: naive B cells (*FCER2^+^TCL1A^+^*), intermediate B cells (*IGHM^+^AIM2^+^*), memory B cells (*IGHM^−^AIM2^+^*), and plasmablasts (*CD38^+^TNFRSF17^+^*) (Figure 3A-C). Only modest changes in their relative proportions were observed following exercise (Figure 3D). However, marked differences in transcriptional responses were detected between B-cell subsets and altitudes. In naive, intermediate, and memory B cells, gene upregulation predominated after low-altitude exercise, whereas gene downregulation predominated after high-altitude exercise. An opposite pattern was observed in plasmablasts (Figure 3E-F).

**Figure 3.**
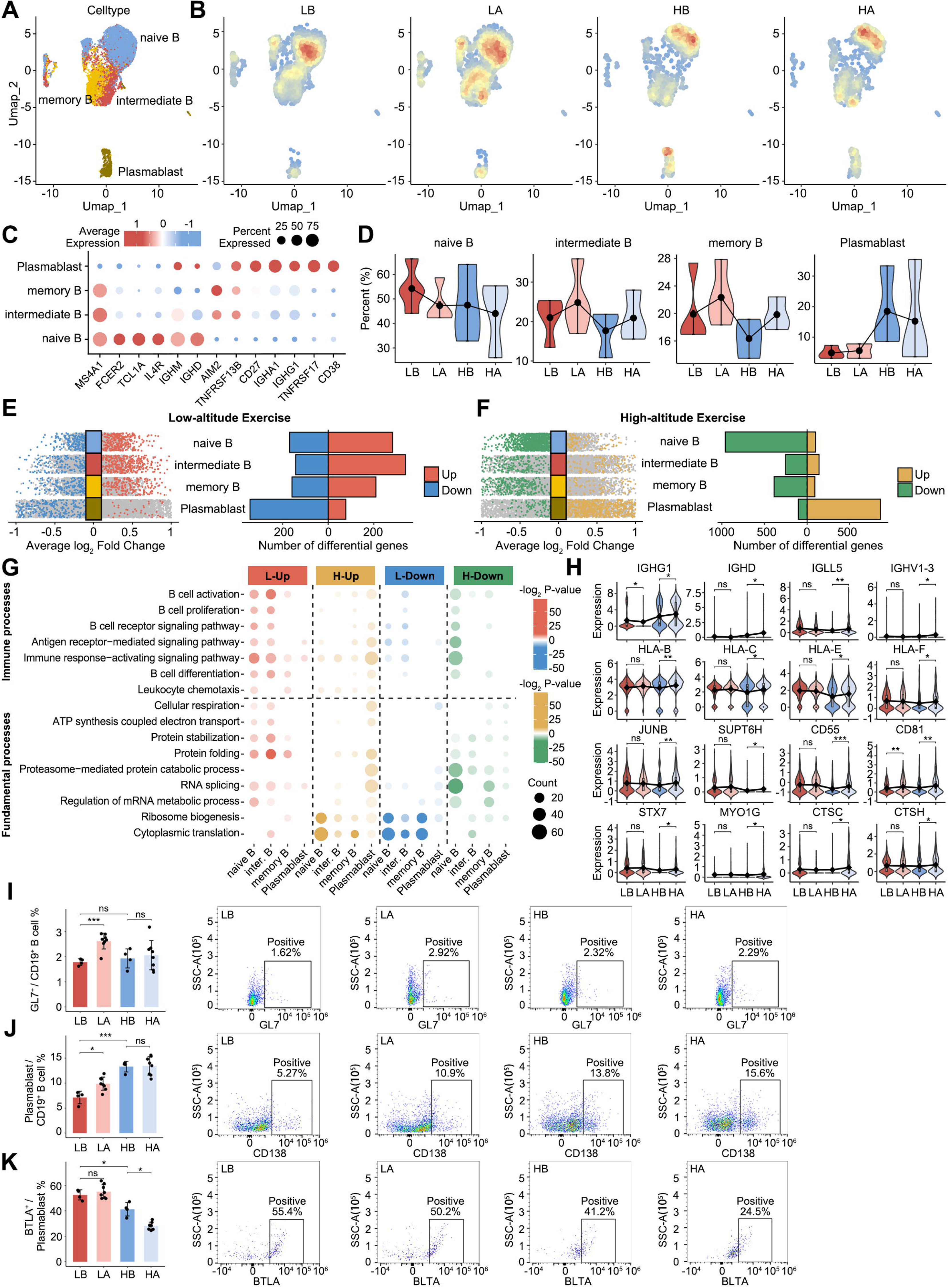
Cellular and functional heterogeneity of B cells in response to exercise at low-altitude and high-altitude. **A.** UMAP plot of the B cell subtypes. **B.** Cell density plots of B cells across four experimental conditions (LB, LA, HB, HA), with color intensity proportional to local cell density. **C.** Dot plots depicting the percentages and average expressions of marker genes in B cell subtypes. **D.** Violin plots showing the changes in the relative proportions of B cell subtypes across four experimental conditions (LB, LA, HB, HA). **E.** Multi-group volcano plots displaying exercise-induced DEGs across B cell subtypes at low-altitude, with significantly upregulated (red) and downregulated (blue) genes indicated (left). Bar plot depicting the number of DEGs in each B cell subtype (right). **F.** Multi-group volcano plots displaying exercise-induced DEGs across B cell subtypes at high-altitude, with significantly upregulated (orange) and downregulated (green) genes indicated (left). Bar plot depicting the number of DEGs in each B cell subtype (right). **G.** Functional enrichment analysis of DEGs across B cell subtypes under low-altitude and high-altitude exercise conditions. The bubble plot displays significantly enriched GO biological processes for four categories: L-Up (red), H-Up (orange), L-Down (blue), and H-Down (green). Color intensity represents -log_2_(P-value), and bubble size indicates the number of genes enriched in each GO term. **H.** Violin plots showing the immune-related genes in plasmablast across four experimental conditions (LB, LA, HB, HA). **I.** Flow cytometry quantification (left) and representative dot plots (right) showing the percentage of activated GL7 cells within the splenic CD19 B cell population across four experimental conditions in the murine model. **J.** Flow cytometry quantification (left) and representative dot plots (right) showing the percentage of CD138 plasmablasts within the splenic CD19 B cell population. **K.** Flow cytometry quantification (left) and representative dot plots (right) showing the percentage of BTLA cells within the plasmablast population. For H-K, statistical significance is indicated as follows: *p < 0.05, ** p < 0.01, *** p < 0.001, ns = not significant. Bar graphs in I-K are presented as mean ± SD.

Following low-altitude exercise, enrichment of B-cell activation, differentiation, B-cell receptor signaling, energy metabolism, and protein-quality-control processes was observed in naive and intermediate B cells, whereas translation- and ribosome-associated processes were reduced. Following high-altitude exercise, several immune- and protein-quality-control pathways were reduced in naive B cells, while protein-synthesis-associated processes were increased. In plasmablasts, enrichment of immune- and protein-processing-associated pathways was observed (Figure 3G). Consistently, after high-altitude exercise, a significant upregulation of immunoglobulin-related genes (e.g., *IGHG1*), multiple HLA/MHC genes, the transcription factor *JUNB*, and secretion-related genes was observed in plasmablasts (Figure 3H).

B-cell phenotypes were additionally assessed in the murine model. Following low-altitude exercise, an increased proportion of GL7^+^ cells within splenic B cells was observed, whereas no corresponding increase was detected after high-altitude exercise (Figure 3I; Supplementary Figure S1). An increased proportion of splenic plasmablasts was observed after chronic hypoxia relative to the low-altitude baseline, whereas no further increase was induced by subsequent high-altitude exercise (Figure 3J). Instead, a significant reduction in surface BTLA expression was detected on plasmablasts following high-altitude exercise (Figure 3K). This reduction in BTLA expression was consistent with the transcriptional profile suggestive of a more activated plasmablast state.

#### 3.2 Effects of Low- and High-Altitude Exercise on CD4 T-Cell Differentiation and Function

Within the CD4^+^ T-cell compartment, six subsets were identified: naive, central-memory (TCM), effector-memory (TEM), cytotoxic, regulatory, and proliferating T cells (Supplementary Figure S2A-C). Only modest shifts in subset proportions were apparent (Supplementary Figure S2D). Naive CD4^+^ T cells, TCM cells, and regulatory T cells showed predominantly upregulated DEGs after low-altitude exercise but predominantly downregulated DEGs after high-altitude exercise. Proliferating CD4^+^ T cells displayed the opposite transcriptional pattern (Supplementary Figure S2E-F).

At low altitude, pathways related to T-cell activation, differentiation, proliferation, chemotaxis, mitochondrial respiration, and protein homeostasis were enriched in naive and memory CD4^+^ T cells. A more selective response was detected after high-altitude exercise. Activation-, chemotaxis-, and differentiation-associated programs were enriched in TCM cells, together with increased *JAK1* and *JUNB* expression, whereas TEM cells showed enrichment of activation- and cytokine-production-associated programs. Protein translation and folding pathways were reduced in both memory populations. Proliferating CD4^+^ T cells, in contrast, showed enrichment of cell-cycle, respiratory, translational, and ribosome-biogenesis processes, accompanied by increased *CCNB2*, *E2F2*, *SBDS*, and *UTP11* expression (Supplementary Figure S2G–I).

Murine flow cytometry showed a higher proportion of CD25^+^ cells within the CD4^+^ TEM compartment after high-altitude exercise (Supplementary Figure S2J; gating strategy in Supplementary Figure S3). PD-1 expression was simultaneously reduced within resting CD25^−^ CD4^+^ TEM cells (Supplementary Figure S2K), while additional changes in CD4^+^ T-cell subsets and TCM phenotypes are shown in Supplementary Figure S4. These results were consistent with selective alteration of memory CD4^+^ T-cell states rather than uniform activation of the CD4^+^ compartment. Taken together, the adaptive immune response to high-altitude exercise was characterized by selective changes in plasmablasts and specific CD4^+^ memory/proliferating populations, while broader activation of naive lymphocyte populations was not evident.

### 4. High-altitude exercise broadly dampens innate and cytotoxic effector lineages spanning CD8^+^ T cells, NK cells, and monocytes/DCs

#### 4.1 Effects of Exercise on CD8^+^ T-Cell Differentiation and Function at Low-altitude and High-altitude

The CD8+ T-cell compartment comprised naive, central-memory, effector-memory, and proliferating populations (Supplementary Figure S5A-C). In naive, effector-memory, and proliferating CD8^+^ T cells, low-altitude exercise was dominated by gene upregulation, whereas high-altitude exercise was dominated by gene downregulation (Supplementary Figure S5E-F).

Immune-related pathways were enriched in naive and effector-memory CD8^+^ T cells after low-altitude exercise but were reduced under high-altitude conditions (Supplementary Figure S5G). The clearest changes in CD8^+^ TEM cells involved reduced expression of genes associated with T-cell receptor signaling (*LCK, ZAP70, CD3D*), cytotoxicity (*KLRD1, KLRC4–KLRK1, NCR3*), and NF-κB signaling (*IKBKB, NFKBIA, MYD88*) (Supplementary Figure S5H).

An increase in CD25^+^ CD8^+^ TCM cells was detected in mice after low-altitude exercise (Supplementary Figure S5I). No significant changes in CD25 expression within CD8^+^ TEM cells were found after high-altitude exercise (Supplementary Figure S5J), and PD-1 expression across CD8^+^ memory compartments also remained unchanged (Supplementary Figure S6). Thus, phenotypic activation comparable with that detected after low-altitude exercise was not evident in the CD8^+^ memory compartment under high-altitude conditions.

#### 4.2 Effects of Low- and High-Altitude Exercise on Monocyte and DC Function

Three monocyte subsets and four DC subsets were identified, including classical, intermediate, and non-classical monocytes, as well as cDC1, cDC2, pDC, and AXL^+^SIGLEC6^+^ DCs (Supplementary Figure S7A-C). Only modest changes in subset proportions were observed after exercise (Supplementary In monocytes, low-altitude exercise was associated with enrichment of innate immune-response and pattern-recognition pathways, whereas these responses were attenuated in classical monocytes after high-altitude exercise (Supplementary Figure S7E-G). DC subsets showed greater heterogeneity. cDC1s were predominantly downregulated after exercise at both altitudes, whereas cDC2s shifted from modest upregulation at low altitude to predominant downregulation at high altitude. Immune-related processes in cDC2s and pDCs were enriched after low-altitude exercise but were generally reduced under high-altitude conditions (Supplementary Figure S7H). In the murine model, no significant exercise-associated changes in monocyted or DC subset proportions were detected. However, CD86 expression was significantly reduced on classical monocytes after high-altitude exercise (Supplementary Figure S7l; Supplementary Figures S8-S9), consistent with the attenuated immune-response programs observed at the transcriptomic level.

#### 4.3 Effects of Low- and High-Altitude Exercise on NK cell Function

To dissect the specific alterations in NK cell subtypes induced by low- and high-altitude exercise, the NK cell lineage was further analyzed. Based on the expression of canonical markers, the NK cell population was classified into five subsets: CD56^bright^ NK, CD56^dim^ KLRG1^+^ NK, CD56^dim^ KLRG1^-^ NK, heat shock protein-associated NK cells (HSP NK), and proliferating NK cells (Figure7A-C). A relatively pronounced decrease in Proliferating NK cells was observed after high-altitude exercise, whereas other subsets exhibited only minor proportional changes post-exercise (Figure 7D). To assess the divergent transcriptional impacts of exercise at different altitudes, differential expression analysis was performed on each NK subset. The results indicated that while CD56^bright^ NK, CD56^dim^ KLRG1 NK, and Proliferating NK cells showed a predominant upregulation of DEGs after low-altitude exercise, all NK subsets exhibited a predominant downregulation after high-altitude exercise (Figure 7E-F).

Enrichment analysis revealed that after low-altitude exercise, immune-related pathways in CD56^bright^ NK and CD56^dim^ KLRG1 NK cells were significantly activated. These included key biological processes such as the regulation of innate immune response, activating signal transduction for immune response, and response to virus. In contrast, after high-altitude exercise, a marked suppression of immune-related pathways was observed in CD56^dim^ NK cells (Figure 7G). In summary, these data suggest that low-altitude exercise broadly activates the immune functions of both CD56^bright^ and CD56^dim^ NK cells. Conversely, high-altitude exercise specifically suppresses the immune and fundamental biological process of CD56^dim^ NK cells, the primary cytotoxic subset^14^.

Taken together, these multi-lineage observations reveal a stark environmental dichotomy in the cellular immune response to acute exercise. While low-altitude exercise broadly promotes the functional activation and immune transcriptional programs of innate and cytotoxic lineages, exhaustive exercise under severe high-altitude hypoxia exerts a coordinated suppressive effect. Specifically, high-altitude exercise broadly downregulates the transcriptional immune and cytotoxicity programs in CD8 T cells and the primary cytotoxic CD56dim NK cells. Concurrently, it significantly reduces CD86 expression on classical monocytes, limiting their capacity to provide crucial co-stimulatory signals. Ultimately, this synchronized dampening of antigen-presenting capacity and cytotoxic effector functions reflects a comprehensive restriction of cellular immune defenses during exhaustive exercise in extreme hypoxic environments.

### 5. High-altitude exercise robustly mobilizes mature neutrophils, driving the specific expansion of an activated phenotype

Neutrophils represented a notable exception to the broadly suppressed transcriptional response observed in other immune lineages after high-altitude exercise. Two neutrophil populations were identified on the basis of canonical markers: mature neutrophils (*CSF3R^+^NAMPT^+^S100A11^+^*) and precursor neutrophils (*CTSC^+^PRSS57^+^HEXA^+^*) (Figure 4A-C). At low altitude, the proportion of precursor neutrophils decreased slightly after exercise, with a corresponding increase in mature neutrophils, whereas the opposite trend was observed at high altitude (Figure 4D). Distinct transcriptional responses were also evident between the two populations. Mature neutrophils showed predominantly upregulated DEGs after exercise at both altitudes, while precursor neutrophils shifted from predominantly upregulated DEGs at low altitude to predominantly downregulated DEGs at high altitude (Figure 4E-F).

**Figure 4.**
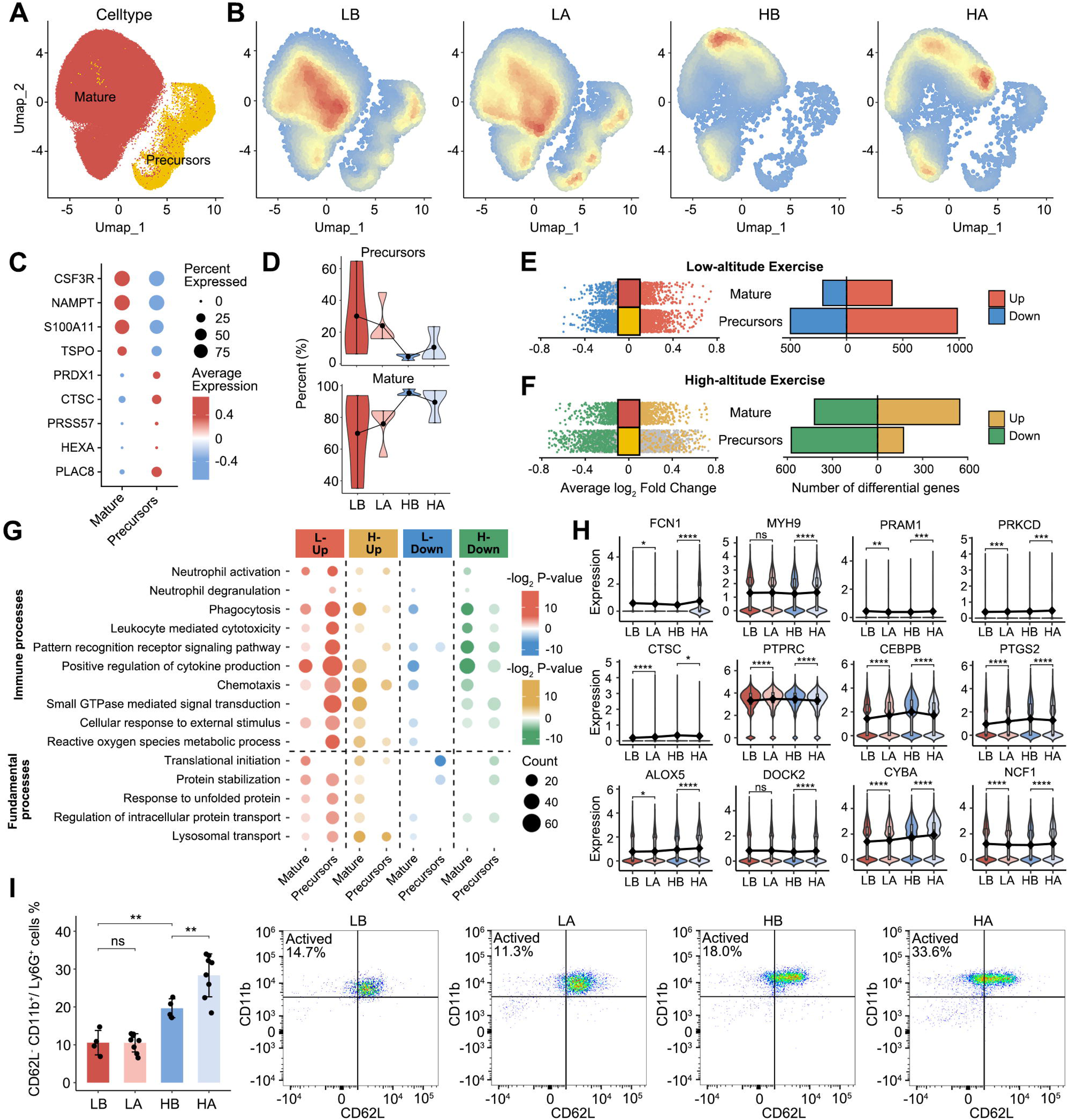
Cellular and functional heterogeneity in the neutrophil in response to exercise at low-altitude and high-altitude. **A.** UMAP plot of the neutrophil subtypes. **B.** Cell density plots of neutrophils across four experimental conditions (LB, LA, HB, HA), with color intensity proportional to local cell density. **C.** Dot plots depicting the percentages and average expressions of marker genes in neutrophil subtypes. **D.** Violin plots showing the changes in the relative proportions of neutrophil subtypes across four experimental conditions (LB, LA, HB, HA). **E.** Multi-group volcano plots displaying exercise-induced DEGs across neutrophil subtypes at low-altitude, with significantly upregulated (red) and downregulated (blue) genes indicated (left). Bar plot depicting the number of DEGs in each neutrophil subtype (right). **F.** Multi-group volcano plots displaying exercise-induced DEGs across neutrophil subtypes at high-altitude, with significantly upregulated (orange) and downregulated (green) genes indicated. Bar plot depicting the number of DEGs in each neutrophil subtype (right). **G.** Functional enrichment analysis of DEGs across neutrophil subtypes under low-altitude and high-altitude exercise conditions. The bubble plot displays significantly enriched GO biological processes for four categories: L-Up (red), H-Up (orange), L-Down (blue), and H-Down (green). Color intensity represents -log_2_(P-value), and bubble size indicates the number of genes enriched in each GO term. **H.** Violin plots showing representative genes involved in chemotaxis (ALOX5, DOCK2), degranulation (PRAM1, PRKCD), phagocytosis (FCN1, MYH9), reactive oxygen species (CYBA, NCF1), cytotoxicity (CTSC, PTPRC), and cytokine production (CEBPB, PTGS2) in mature neutrophils across four experimental conditions (LB, LA, HB, HA). **I.** Flow cytometry quantification (left) and representative dot plots (right) showing the percentage of activated mature neutrophils (phenotypically defined as CD62L CD11b) within the peripheral blood Ly6G neutrophil population across four experimental conditions in the murine model. For H and I, statistical significance is indicated as follows: *p < 0.05, **p < 0.01, ***p < 0.001, ****P < 0.0001, ns = not significant. The bar graph in I is presented as mean ± SD.

Functional enrichment further revealed marked differences between altitude conditions. At low altitude, exercise-related changes were concentrated mainly in precursor neutrophils, with enrichment of neutrophil activation, chemotaxis, pattern-recognition, and protein-quality-control pathways, whereas comparatively limited changes were detected in mature neutrophils. In contrast, high-altitude exercise was associated with a distinct transcriptional state in mature neutrophils. Chemotaxis, responses to external stimuli, reactive oxygen species metabolism, and several protein-processing pathways were enriched, while phagocytosis- and degranulation-associated programs were maintained. At the same time, cytotoxicity-, pattern-recognition-, and cytokine-production-associated pathways were reduced (Figure 4G-H). These results indicate that mature neutrophils retain prominent migratory and stress-response programs after high-altitude exercise despite attenuation of several inflammatory effector pathways.

A related change in circulating neutrophil phenotype was identified in the murine model. The proportion of CD62L^−^CD11b^+^ cells within peripheral blood Ly6G^+^ neutrophils increased significantly after high-altitude exercise, whereas no significant change was detected following low-altitude exercise (Figure 4I; Supplementary Figure S11). Thus, high-altitude exercise was characterized by a distinct mature-neutrophil transcriptional response in humans and an increased circulating CD62L^−^CD11b^+^ neutrophil population in mice.

### 6. Systemic proteomic and skeletal muscle transcriptomic profiling reveals a high-altitude-specific vascular extravasation program

Plasma proteomic profiling of 40 participants identified 4,461 proteins before and after high-altitude exercise. Among them, 115 were significantly increased and 31 decreased after exercise at an FDR < 0.05 (Figure 5A; Supplementary Table 4). Separation of pre- and post-exercise samples was evident by principal component analysis based on these differentially expressed proteins (Figure 5B).

**Figure 5.**
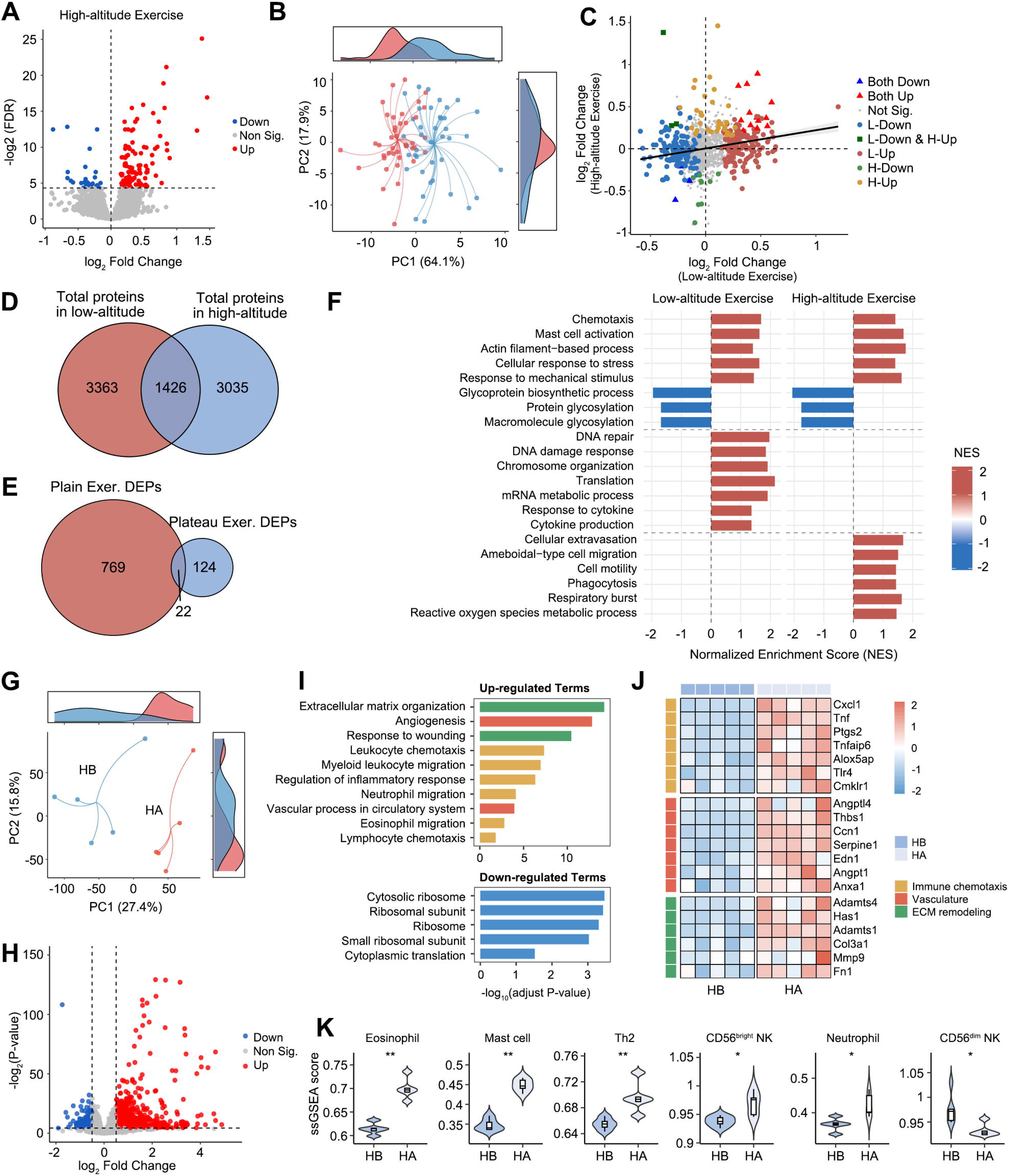
Untargeted proteomics reveals exercise-induced plasma proteome alterations at low-altitude and high-altitude. **A.** Volcano plot displaying DEPs following high-altitude exercise, with significantly upregulated (red) and downregulated (blue) proteins indicated. Gray dots represent non-significant changes. **B.** Principal component analysis (PCA) showing the separation of plasma proteome profiles between HB (blue) and HA (red) samples at high-altitude. Density distributions along PC1 and PC2 axes are shown at the margins. **C.** Scatter plot comparing log_2_ fold changes of proteins between low-altitude exercise (x-axis) and high-altitude exercise (y-axis). Colors indicate different regulation patterns: proteins down-regulated in both conditions (Both Down), up-regulated in both conditions (Both Up), non-significant changes (Not Sig.), and altitude-specific changes including low-altitude-specific down-regulation (L-Down), high-altitude-specific down-regulation (H-Down), low-altitude-specific up-regulation (L-Up), high-altitude-specific up-regulation (H-Up), and opposite regulation pattern (L-Down & H-Up). **D.** Venn diagram showing the overlap of total detected proteins between low-altitude exercise (4,789 proteins, red) and high-altitude exercise (4,461 proteins, blue), with 1,426 proteins detected in both conditions. **E.** Venn diagram illustrating the overlap of DEPs between low-altitude exercise (791 DEPs, red) and high-altitude exercise (146 DEPs, blue), with 22 common DEPs. **F.** Bar plot displaying normalized enrichment scores (NES) of selected GO biological processes from GSEA in low-altitude exercise and high-altitude exercise. **G.** PCA showing the separation of skeletal muscle transcriptome profiles between HB (blue) and HA (red) samples. Density distributions along PC1 and PC2 axes are shown at the margins. **H.** Volcano plot of DEGs in skeletal muscle following high-altitude exercise, with significantly upregulated (red) and downregulated (blue) genes indicated. Gray dots represent non-significant changes. **I.** GO biological process enrichment analysis of upregulated (upper panel) and downregulated (lower panel) DEGs. Bar colors correspond to functional categories as in panel J. **J.** Heatmap showing Z-score normalized expression of representative upregulated genes across three enriched functional categories: immune chemotaxis (yellow), vasculature (red), and ECM remodeling (green). Each column represents an individual sample. **K.** ssGSEA enrichment scores of immune cell populations showing significant differences between HB and HA groups. For K, statistical significance was determined by the Wilcoxon rank-sum test: *p < 0.05, **p < 0.01.

Comparison with a published low-altitude exercise proteomic dataset revealed limited concordance between the two exercise responses. Protein fold changes were only weakly correlated (R = 0.209) (Figure 5C), and among 1,426 proteins detected in both datasets, only 22 DEPs were shared (Figure 5D-E).

GSEA identified a group of responses common to both exercise conditions, including chemotaxis, locomotion, actin-filament-associated processes, mast-cell activation, and responses to stress and mechanical stimuli (Figure 5F). Low-altitude exercise additionally showed enrichment of DNA repair, chromosome organization, translation, mRNA metabolism, and cytokine-associated processes. By contrast, the cellular-extravasation gene set was significantly enriched only after high-altitude exercise (NES = 1.68, p = 0.01; low altitude: NES = 0.73, p = 0.87). Ameboid cell migration, cell motility, phagocytosis, respiratory burst, and reactive oxygen species metabolism were also preferentially enriched in the high-altitude dataset.

A related pattern was detected in skeletal muscle from the murine model. RNA sequencing identified 957 upregulated and 228 downregulated genes after high-altitude exercise (Figure 5G-H). The upregulated genes were enriched in leukocyte and neutrophil migration, angiogenesis, and vascular-permeability-related processes (Figure 5I). Increased expression of Cxcl1, Tnf, Thbs1, and Serpine1 was also detected (Figure 5J). In parallel, ssGSEA scores for neutrophils, mast cells, and eosinophils were higher after exercise (Figure 5K). Taken together, the plasma and skeletal-muscle datasets converged on a chemotactic and vascular-response signature associated with high-altitude exercise and compatible with increased innate immune-cell recruitment to skeletal muscle.

### 7. Integrated transcriptomic-proteomic analysis reveals exercise-induced remodeling of plasma-immune cell communication networks at high-altitude

Integration of plasma proteomic data with cell-type-specific receptor expression revealed substantial differences in ligand–receptor communication after high-altitude exercise. In the global communication network, monocytes and neutrophils received the strongest signaling inputs, whereas communication toward adaptive lymphocytes was comparatively limited (Supplementary Figure S12A). At the pathway level, communication strength increased for ANNEXIN, APRIL, and MIF signaling, while CCL and COMPLEMENT signaling decreased after exercise (Supplementary Figure S12B-C).

Within the APRIL pathway, plasma TNFSF13 was increased after exercise, accompanied by an upward trend in TNFRSF13B expression in plasmablasts (Supplementary Figure S12D, I). In contrast, attenuation of CCL signaling was accompanied by reduced plasma CCL14 and decreased expression of CCR1 and CCR2 in monocytes (Supplementary Figure S12E, J). A similar pattern was observed for COMPLEMENT signaling, with lower plasma C3 together with reduced ITGAM and ITGB2 expression in NK cells and monocytes (Supplementary Figure S12F, K).

Distinct changes were also identified in signaling pathways involving neutrophils. Plasma MIF increased markedly after high-altitude exercise, whereas CXCR2 expression in mature neutrophils decreased; the overall MIF communication strength nevertheless increased (Supplementary Figure S12G, L). In the ANNEXIN pathway, increased plasma ANXA1 was accompanied by increased FPR2 expression in neutrophils and a corresponding increase in pathway-level communication strength (Supplementary Figure S12H, M). Collectively, high-altitude exercise was associated with coordinated remodeling of plasma-immune-cell communication, characterized by enhanced APRIL, MIF, and ANNEXIN signaling together with attenuation of CCL and COMPLEMENT pathways.

## Discussion

This study provides a comprehensive, multi-omics map of the immune system’s dynamic response to acute exercise at both low-altitude and high-altitude, based on an integrated analysis of plasma proteomics and single-cell transcriptomics. Through the integration of these datasets, a unique immune dysregulation characteristic of high-altitude exercise was successfully captured. A two-pronged response was revealed: on one hand, neutrophils were found to undergo metabolic reprogramming and stress-induced degranulation to establish a rapid defense barrier ^15^; on the other, an imbalance emerged between cellular and humoral immunity. This was characterized by a widespread suppression of cellular immunity mediated by CD8 T cells and NK cells, juxtaposed with a partial activation of humoral immunity driven by CD4 TCM cells and plasmablasts. This immunoregulatory pattern unveils a unique adaptive mechanism of the human body to the dual stressors of hypoxia and exercise.

Prior to this study, progress in characterizing these complex immune responses had been hampered by inherent methodological limitations. Previous investigations were typically confined to observing resting states at high-altitude or exercise responses at sea level, or were conducted during uncontrolled mountaineering expeditions where numerous variables were confounded^8, 12, 16, 17^. Furthermore, studies relying on bulk transcriptomics could only detect the average signal across mixed cell populations. Such bulk-level analyses inherently mask potentially divergent or even opposing responses among distinct immune cell subsets—an omission that obscures the cell-type-specific imbalances critical to understanding the overall immunological shift^18^. The within-participant longitudinal design, together with standardized exercise conditions and consistent dietary and habitual physical-activity conditions, reduced inter-individual variability and several potential behavioral confounders. Nevertheless, because altitude exposure necessarily followed the low-altitude assessment, the design cannot completely separate altitude effects from other time-dependent factors.

Neutrophils, as the primary effector cells of the innate immune system, are rapidly recruited to sites of infection or injury, where pathogens are cleared via phagocytosis, degranulation, and the formation of Neutrophil Extracellular Traps (NETs)^19^. It is well-established that the effect of exercise on neutrophils is intensity-dependent^20^; high-intensity exercise rapidly mobilizes neutrophils from the bone marrow and marginated pools into circulation, leading to a significant increase in peripheral white blood cell count^21^. This process, primarily driven by catecholamines and cortisol, not only promotes cell release but also enhances their adhesion and migration capabilities^22, 23^. The findings from the low-altitude exercise condition in this study are consistent with this, as a decrease in the proportion of precursor neutrophils and a corresponding increase in mature neutrophils were observed, reflecting the mobilization of mature cells. More importantly, it is suggested by our results that low-altitude exercise may primarily enhance immune potential by activating the more plastic precursor neutrophils in preparation for potential injury or infection, while mature neutrophils are likely responsible for executing immediate effector functions.

However, the high-altitude hypoxic environment injects new complexity into the immune response of neutrophils. Studies have shown that hypoxia stabilizes HIF-1α, which drives cellular metabolism from oxidative phosphorylation to anaerobic glycolysis—a shift essential for maintaining neutrophil survival, motility, and bactericidal capacity under hypoxic stress^24–26^. Notably, our previous study also observed that chronic high-altitude exposure significantly promoted the differentiation of peripheral blood neutrophils to the terminal maturation stage, accompanied by the baseline activation of granule synthesis and chemotactic effector functions.

In the present study, an in-depth multi-omics analysis revealed exactly how this hypoxic baseline state shifts in response to acute altitude exercise. Rather than a global hyperactivation, the superimposition of exhaustive exercise triggered a highly selective transcriptional reprogramming within mature neutrophils. Single-cell transcriptome data indicated that while chemotaxis and stress responses (e.g., ROS metabolism) were markedly enhanced, conventional cytotoxicity and pro-inflammatory cytokine production were profoundly suppressed. This functional shift was phenotypically accompanied by a robust expansion of activated mature neutrophils (CD62L CD11b) in the peripheral blood. Furthermore, integrated upstream signaling analysis revealed that this reprogramming coincided with the enhancement of a dual-signal communication network. Specifically, plasma MIF was significantly elevated, providing a potential systemic cue for the chemotactic mobilization and subsequent vascular extravasation of neutrophils into skeletal muscle. Concurrently, the Annexin A1 (ANXA1) pathway was strongly upregulated, which likely acts as a vital anti-inflammatory brake^27–29^. Taken together, under extreme hypoxic stress, although neutrophils maintain robust migratory and mobilization activity, their overt cytotoxicity is strictly restrained. This suggests that the body carefully regulates immune resources, seeking a precise balance between targeted activation and protective inhibition to avoid functional exhaustion and tissue damage in an energy-limited environment.

Beyond the innate immune response, acute exercise acts as a potent physiological stressor that significantly alters the distribution and function of adaptive immune cells^30^. Previous studies have shown that vigorous exercise promotes the migration of lymphocytes from the marginal pool and bone marrow into the peripheral blood, and the mobilization pattern of B cells differs from that of T and NK cells. Specifically, acute vigorous exercise primarily mobilizes immature B cells into the peripheral circulation rather than those in an effector state^31^. In line with this, our low-altitude acute exercise experiment showed that the immune activation, differentiation and energy metabolism pathways were significantly enhanced, while RNA translation and ribosome production were down-regulated in naive and intermediate B cells after exercise. This suggests that these B cells are efficiently transitioning to the mature stage, preparing for future immune responses, while maintaining protein quality control. In addition, we did not observe any significant activation of plasmablastic related genes by low-altitude exercise, and only a few molecules such as CD81 were slightly up-regulated, further confirming that the focus of B cell mobilization during acute exercise under low-altitude conditions was the enhancement of reserve and differentiation potential, rather than the immediate antibody secretion.

Chronic high-altitude exposure, however, establishes a globally suppressive environment for adaptive immunity. While moderate hypoxia may transiently prompt B cell differentiation^32^, severe or prolonged hypoxia ultimately restricts class switching and stalls B cells at the memory stage, impairing antibody affinity maturation^33^. Consistent with this, our previous observations confirmed that long-term high-altitude exposure induces an "innate activation-adaptive suppression" pattern, characterized by restricted plasmablast maturation and compromised humoral defense against pathogens. Remarkably, our results revealed that the superimposition of acute exercise on this chronic hypoxic baseline triggered a rapid shift toward a "plasmablast-centric" compensatory mechanism within the humoral immune response. Under this dual stress, a striking functional dichotomy emerged among B cell subsets. In naive, intermediate, and memory B cells, classical immune activation pathways were broadly downregulated, whereas fundamental protein translation and ribosome biogenesis were selectively maintained. This likely reflects a strict energy-conservation strategy under hypoxia, whereby resting B cells reduce their basal responses to preserve synthetic capacity for future demands. Simultaneously, the plasmablast subset exhibited profound compensatory activation. Following high-altitude exercise, genes encoding immunoglobulins and secretion/lysosomal machinery were significantly upregulated in these effector cells, indicating a robust drive for immediate antibody synthesis. This targeted activation suggests that during acute physical stress under extreme hypoxia, the immune system maximizes the secretory function of existing plasmablasts to sustain immediate humoral defense, thereby compensating for the restricted differentiation of immature B cells. Nevertheless, while this emergency activation maintains short-term antibody production, its long-term sustainability and ultimate impact on antibody affinity maturation and memory B cell formation warrant further in-depth investigation.

In conclusion, through the integration of plasma proteomics and single-cell transcriptomics, the first comprehensive map of the immune system’s dynamic response to acute exercise under both low-altitude and high-altitude conditions is provided by this study. It is revealed by our findings how the human immune system navigates the dual challenges of hypoxia and exercise, deepening our understanding of high-altitude physiological adaptation and providing a new theoretical basis for the early warning and intervention of altitude-related health issues. However, this study has limitations. First, the sample size is relatively limited, which may affect the generalizability of the results. Second, the focus was on the immediate post-exercise response, precluding a full understanding of the long-term dynamics. Furthermore, while a comprehensive molecular map is provided by the multi-omics data, causal relationships require further functional validation. Future research should involve larger, longitudinal cohorts to track the long-term evolution of immune adaptation. In-depth mechanistic studies, using in vitro validation and animal models, are needed to dissect the regulatory networks in key cell subsets. Finally, exploring the association between these immunoregulatory patterns and clinical outcomes, such as susceptibility to infection and vaccine efficacy, will provide a solid foundation for developing precise health protection strategies and enhancing human resilience in extreme environments.

## Methods

### Sample collection and ethics statement

The study was conducted at Jiangjin, Chongqing (200 m above sea level) and Lhasa, Tibet (3,650 m above sea level), representing low- and high-altitude environments, respectively. A total of 46 healthy male Chinese volunteers residing at low altitude (<300 m) and without a history of chronic disease were recruited. Participants were instructed to maintain consistent dietary intake and habitual physical activity throughout the study period.

Physiological assessments and fasting venous blood samples were available from 45 participants at low altitude. Participants completed the Cooper 12-min run test on a standard 400-m track following a 10-min warm-up^34^. Blood samples were collected before and after exercise according to the study protocol. After relocation to Lhasa, 40 participants completed the corresponding assessment after 90 days of high-altitude acclimatization, using the same exercise and sampling procedures. These 40 participants were included in the high-altitude proteomic analysis using paired pre- and post-exercise samples.

For single-cell RNA sequencing, four participants were randomly selected from the original cohort and sampled before and after exercise at low altitude. Three of these participants completed repeated sampling after high-altitude acclimatization and were included in the high-altitude scRNA-seq analysis. The study was approved by the Institute Research Medical Ethics Committee of the Army Medical University, and written informed consent was obtained from all participants.

### Proteomic profiling and data processing

Paired pre- and post-exercise samples from the 40 participants who completed the high-altitude assessment were subjected to quantitative proteomic profiling. Samples were processed using a low-abundance protein enrichment workflow and analyzed by data-independent acquisition mass spectrometry on an Orbitrap Exploris 480 platform. Raw data were processed using Spectronaut 18 (Biognosys AG) in library-free Direct-DIA mode against the human reference proteome. Detailed sample preparation, chromatographic conditions, mass-spectrometry acquisition settings, and database-search parameters are provided in Supplementary Methods.

Missing abundance values were imputed using the k-nearest-neighbor algorithm, followed by median normalization. Differential protein abundance between pre- and post-exercise samples was assessed using paired Student’s t-tests with Benjamini-Hochberg correction, and proteins with an FDR <0.05 were considered differentially expressed.

To compare exercise-induced proteomic responses between altitudes, the high-altitude dataset was further compared with a published low-altitude exercise plasma proteomic dataset^35^. Proteins detected in both datasets were retained for comparative analysis. Gene set enrichment analysis was performed using Gene Ontology Biological Process gene sets, with 1,000 permutations and nominal P<0.05 used as the enrichment threshold.

### Single-cell sequencing and data processing

Sample preparation, scRNA-seq library construction, quality control, dimensionality reduction, batch correction, clustering, and cell-type annotation were performed according to the previously established pipeline^36^. Detailed procedures are provided in Supplementary Methods.

Exercise-associated transcriptional changes were analyzed separately within each cell type and subtype at low and high altitude using the FindMarkers function in Seurat. Differentially expressed genes were identified using the Wilcoxon rank-sum test with |log_2_ fold change| ≥ 0.1 and adjusted P < 0.05. Functional enrichment analysis was performed using clusterProfiler (v4.10.0).

To identify shared transcriptional patterns across immune-cell lineages, genes showing concordant exercise-associated changes in multiple major immune-cell populations were subjected to fuzzy c-means clustering. Pseudo-bulk expression profiles were generated across the four experimental conditions (LB, LA, HB, and HA), and six expression clusters were identified. Gene-selection criteria, clustering parameters, and enrichment procedures are described in Supplementary Methods.

### Integrated transcriptomic- Proteomic pathway analysis

A cross-omics analysis was performed to evaluate changes in potential communication between circulating proteins and peripheral immune cells following high-altitude exercise. Ligand–receptor interactions were obtained from the CellChat database. Proteins detected by proteomic profiling were treated as potential circulating ligands, whereas receptor genes expressed in scRNA-seq-defined immune-cell populations were treated as potential receptors.

Ligand and receptor abundance was summarized using a quantile-based expression metric, from which ligand–receptor and pathway-level communication scores were calculated. Exercise-associated changes were determined by comparing pre- and post-exercise communication scores. The complete calculation procedure, normalization strategy, and equations are provided in Supplementary Methods.

### *In Vivo* Murine Model and Exercise Protocol

Male BALB/c mice aged 6–8 weeks were randomly assigned to four experimental groups (n=8 per group): low-altitude baseline (LB), low-altitude exercise (LA), high-altitude baseline (HB), and high-altitude exercise (HA). Mice in the HB and HA groups were continuously exposed for 44 days to hypobaric hypoxia simulating an altitude of 5,800 m (approximately 50 kPa).

Following hypoxic exposure, mice in the LA and HA groups underwent the same exhaustive treadmill exercise protocol. The treadmill was set at a 10° incline, with a 5-min warm-up at 10 m/min followed by an increase in speed of 1.5 m/min every minute to a maximum of 40 m/min. Exhaustion was defined as remaining on the shock grid for >10 consecutive seconds without attempting to resume running.

Peripheral blood and spleens were collected for flow-cytometric analysis. Flow cytometry included eight mice in each exercise group and four mice in each corresponding baseline group. Gastrocnemius muscle from HB and HA mice (n=5 per group) was collected for bulk RNA sequencing. All animal experiments were approved by the Institute Research Medical Ethics Committee of the Army Medical University.

### Murine Sample Preparation and Flow Cytometry

Peripheral blood and spleens were collected immediately after exhaustive exercise or at matched resting time points. Single-cell suspensions were prepared, erythrocytes were lysed, and viable cells were stained with fluorochrome-conjugated antibodies.

Flow-cytometric panels were used to characterize circulating neutrophils and splenic B-cell, T-cell, monocyte, and dendritic-cell populations and their selected phenotypic markers. Data were acquired using a FongCyte S four-laser flow cytometer (SinoCyte) and analyzed using FlowJo. Antibody panels, catalogue numbers, and gating strategies are provided in Supplementary Methods and the corresponding Supplementary Figures.

### Muscle bulk RNA sequencing and data processing

Gastrocnemius muscle from HB and HA mice (n=5 per group) was subjected to bulk RNA sequencing. Poly(A)-enriched libraries were sequenced on an Illumina platform using paired-end 150-bp reads. After quality control and trimming, reads were aligned to the mouse reference genome (GRCm38/mm10), and gene-level counts were generated for downstream analysis. Detailed RNA extraction, library-preparation, sequencing, and alignment procedures are provided in Supplementary Methods.

Differential expression between HA and HB groups was analyzed using DESeq2 (v1.42.0). Genes with P < 0.05 and |log_2_ fold change| > 0.5 were considered differentially expressed, followed by Gene Ontology enrichment analysis using clusterProfiler.

Immune-cell-associated signatures in skeletal muscle were estimated by single-sample gene set enrichment analysis (ssGSEA) using GSVA (v1.50.5) and previously published immune-cell marker gene sets^37^. Differences between HB and HA groups were assessed using the Wilcoxon rank-sum test.

### Statistics and reproducibility

Statistical analyses were performed using R (v4.1.1). Statistical tests specific to individual omics analyses are described above. Unless otherwise specified, two-tailed Wilcoxon rank-sum tests were used for group comparisons. Proteomic pre- versus post-exercise comparisons were analyzed using paired Student’s t-tests followed by Benjamini–Hochberg correction. Statistical significance was defined as P<0.05. Sample sizes and statistical tests for individual experiments are additionally indicated in the corresponding figure legends.

## Supplementary Material

Document S1. Supplementary Methods and Figures S1-S12.

Table S1. Effects of acute exercise on physiological parameters at low- and high-altitude.

Table S2. Volunteer Information and Physiological Data.

Table S3. Differentially expressed genes in peripheral blood cell types and subsets.

Table S4. Differential plasma proteomic analysis in response to high-altitude exercise.

Table S5. Differentially expressed genes in skeletal muscle following high-altitude exercise.

## Acknowledgements

We sincerely thank the 46 volunteers who enthusiastically participated in this work. We acknowledge the General Hospital of Tibet Military Area Command for their invaluable assistance. This work was financially supported by the Noncommunicable Chronic Diseases-National Science and Technology Major Project (Grant No. 2025ZD0551904) and the Young Scientists Fund of the National Natural Science Foundation of China (Grant No. 82502270).

## Author contributions statement

J.L.Z., Y.P.T., Y.R. and Yan H. designed the project. J.Q.W., R.Y.X., Z.J.T. and W.B.Y. performed the bioinformatics analysis. J.L.Z., Y.T.D., C.Z., Y.K.Z., G.X.H.Z., M.Y.G., R.J. and Yi H. performed the experiments. J.L.Z., J.Q.W. and R.Y.X. wrote the manuscript. Y.P.T., Y.R. and Yan H. conceived and supervised the study. J.L.Z., J.Q.W., Y.R. and Yan H. contributed to the analysis and interpretation of data.

## Data Availability Statement

The single-cell RNA sequencing (scRNA-seq) generated in this study have been deposited in the China National GeneBank (CNGB) Sequence Archive (CNSA; https://db.cngb.org/cnsa/) under accession numbers CNP0008552 (scRNA-seq). The bulk RNA sequencing data of muscle tissues have been deposited in the NCBI Gene Expression Omnibus (GEO) database under accession number GSE337978. The mass spectrometry proteomics data have been deposited to the ProteomeXchange Consortium (https://proteomecentral.proteomexchange.org) via the iProX partner repository^38, 39^ with the dataset identifier PXD071611.

All original analysis code has been made freely accessible at https://github.com/Jack123-Wang/High-Altitude-Exercise-Ruan.

