## Supplementary Material for "High-altitude exercise orchestrates a divergent immune landscape: cytotoxic suppression, humoral compensation, and neutrophil functional reprogramming"

**Single-Cell Transcriptomics and Proteomics Reveal a Divergent Immune Landscape of Acute Exercise at High- Versus Low-Altitude**

Junlei Zhang^1,#^, Jiaqi Wang^1,#^, Ruoyi Xue^2,#^, Yutong Dong^1,3^, Zujie Tang^3^, Chen Zhang^1^, Wubin Yang^1,4^, Yangkai Zhang^1^, Guangxinghao Zhang^1,5^, Manying Guo^2^, Rui Jian^1^, Yi Huang^6^, Yanping Tian^1,*^, Yan Ruan^1,*^ & Yan Hu^3,*^

^1^Laboratory of Stem Cell & Developmental Biology, Department of Histology and Embryology, College of Basic Medical Sciences, Army Medical University, Chongqing 400038, China.

^2^Clinical Laboratory and Department of Pathology, The 72nd Army Hospital of the People's Liberation Army, Huzhou University, Huzhou 313099, PR China.

^3^Department of Military Joint and Force Management, Army Training Base for Health Care, Army Medical University, Chongqing 400038, China.

^4^Department of Pathophysiology, College of High Altitude Military Medicine, Army Medical University, Chongqing 400038, China.

^5^Institute of Neuroscience, School of Basic Medical Sciences, Chongqing Medical University, Chongqing 400016, China.

^6^Biomedical Analysis Center, Army Medical University, Chongqing 400038, China.

^#^These authors contributed equally to this work.

**Supplementary Methods**

**Human exercise protocol**

At the low-altitude site, participants completed the Cooper 12-min run test on a standard 400-m track following a 10-min warm-up. Participants were instructed to cover the greatest possible distance within 12 min, and the total distance was recorded. Maximal oxygen uptake was estimated using the following equation: VO₂max (mL·kg⁻¹·min⁻¹) = 21.01 × distance (km) − 11.04. Heart rate during exercise was maintained between 120 and 180 beats/min. Following 90 days of acclimatization in Lhasa (3,650 m), the same exercise and sampling procedures were repeated.

**Proteomic sample preparation and LC–MS/MS analysis**

Samples were processed using a Low-abundance Protein Enrichment Kit (OMIC SOLUTION, OSFP0002) according to the manufacturer's instructions. Briefly, 100 μL of sample was incubated with pre-washed magnetic beads. Following depletion of high-abundance proteins, retained proteins were subjected to on-bead enzymatic digestion for 2 h. Peptides were purified using solid-phase extraction and subsequently lyophilized.

Dried peptides were reconstituted in 0.1% formic acid and analyzed using an UltiMate 3000 RSLCnano system (Thermo Fisher Scientific) equipped with a 25-cm nano-electrospray integrated analytical column (IonOpticks, AUR3-25075C18). Separation was performed at a flow rate of 300 nL/min using solvent A (0.1% formic acid in water) and solvent B (0.1% formic acid in 80% acetonitrile). The gradient was 5–20% B over 2 min, 10–40% B over 80 min, 40–55% B over 2 min, 55–90% B over 2 min, and 90–100% B over 5 min.

Peptides were introduced into an Orbitrap Exploris 480 mass spectrometer (Thermo Fisher Scientific) by nano-electrospray ionization at 2.5 kV. Data were acquired in data-independent acquisition mode. Full MS scans were collected over m/z 350–1200 at a resolution of 120,000, and MS/MS spectra were acquired at a resolution of 30,000 using 42 variable isolation windows.

Raw Direct-DIA data were processed using Spectronaut 18 (Biognosys AG) in library-free mode against the human UniProt reference proteome. Trypsin/P was specified as the protease, allowing up to two missed cleavages. Carbamidomethylation of cysteine was set as a fixed modification, whereas methionine oxidation and N-terminal acetylation were treated as variable modifications. FDR thresholds were maintained at 1% for peptide-spectrum matches and peptides and 5% for proteins.

Missing protein-abundance values were imputed using the k-nearest-neighbor algorithm, and protein-abundance values were median-normalized across samples. Differential abundance was assessed using paired Student's t-tests, with P values adjusted using the Benjamini–Hochberg method. Proteins with FDR <0.05 were considered differentially expressed.

For comparison with the published low-altitude exercise proteomic dataset, proteins detectable in both datasets were retained. Log2 fold-change values were used to generate ranked gene lists for GSEA. Gene Ontology Biological Process gene sets were obtained from MSigDB, and GSEA was performed independently for each dataset using 1,000 permutations. Pathways with nominal P < 0.05 were considered significantly enriched.

**Single-cell RNA sequencing and bioinformatic analysis**

For each defined cell type or subtype, exercise-associated differential expression was assessed separately at low and high altitude using the FindMarkers function in Seurat. Post-exercise samples were compared with the corresponding baseline samples using the Wilcoxon rank-sum test. DEGs were defined by |log_2_FC| ≥ 0.1 and adjusted P < 0.05. Complete DEG lists are provided in Supplementary Table 3.

For identification of shared transcriptional trajectories, genes with P < 0.05 showing concordant directional changes in at least four of seven major immune-cell populations—B cells, CD4^+^ T cells, CD8^+^ T cells, dendritic cells, monocytes, neutrophils, and NK cells—were selected, with platelet-associated genes excluded. Pseudo-bulk expression values across LB, LA, HB, and HA were calculated using AverageExpression in Seurat.

Fuzzy c-means clustering was performed using the Mfuzz algorithm implemented in ClusterGVis (v1.0.0). Genes were assigned to six clusters on the basis of their expression trajectories, and genes with membership values > 0.35 were retained. Gene Ontology Biological Process enrichment was performed using enrichCluster with adjusted P < 0.05. Cell-type-specific functional enrichment of DEGs was performed using clusterProfiler (v4.10.0).

**Integrated proteomic–single-cell ligand–receptor analysis**

To investigate the changes in signaling communication between plasma proteins and peripheral blood immune cells in response to high altitude exercise, we developed an integrated analytical approach combining plasma proteomics and single-cell transcriptomics.

*Selection of Ligand-Receptor Pairs*

Ligand-receptor interaction information from the CellChat database was used as reference. Proteins detected in plasma proteomics were considered as potential ligands, while genes expressed in immune cell subpopulations identified by single-cell RNA sequencing were considered as potential receptors. Ligand-receptor pairs for downstream analysis were determined by intersecting detected molecules with the CellChat database.

*Calculation of Expression Metric (EM)*

Referring to the communication strength calculation method of CellChat with modifications, we employed a quantile-based weighted expression metric to quantify the expression levels of ligands and receptors. The metric was calculated as follows:

$$EM=0.4\times Q_{2}+0.3\times(Q_{1}+Q_{3})+0.3\times(P_{10}+P_{90})$$

where*Q_1_*, *Q_2_* and *Q_3_* represent the 25th, 50th, and 75th percentiles, respectively, and *P_10_* and *P_90_* represent the 10th and 90th percentiles, respectively. Compared to the original CellChat method, the *P_10_* and *P_90_* terms were introduced to reduce computational bias caused by expression values fluctuating around quantile thresholds.

For plasma protein ligands, *EM* values were calculated separately for pre-exercise and post-exercise samples. For receptor genes, *EM* values were calculated by cell type and time point. Subsequently, *EM* values for ligands and receptors were scaled separately by dividing by their respective standard deviations without mean centering.

$${EM}_{scaled}=\frac{EM}{SD(EM)}$$

where *EM_scaled_* represents the standardized expression metric, *EM* is the original expression metric calculated above, and *SD(EM)* is the standard deviation of *EM* values. This scaling was performed independently for ligands and receptors by dividing by their respective standard deviations without mean centering. In all subsequent calculations, *EM* refers to the standardized expression metric (*EM_scaled_*).

*Calculation of Ligand-Receptor Communication Strength*

The Ligand-Receptor Communication Score (LRCS) was used to quantify the communication strength between ligands and receptors. For single-subunit receptors, LRCS was defined as the product of ligand EM and receptor EM:

$$LRCS={EM}_{ligand} \times{EM}_{receptor}$$

For heterodimeric receptors composed of two subunits, the geometric mean of the two subunit EM values was first calculated as the composite receptor expression metric, which was then multiplied by the ligand EM:

$${EM}_{receptor}=\sqrt{{EM}_{R1}\times{EM}_{R2}}$$

*Calculation of Signaling Pathway-Level Communication Strength*

When a signaling pathway contained multiple ligand-receptor pairs, the relative contribution weight of each pair within the pathway was calculated based on their LRCS values, and the pathway-level weighted communication score was subsequently derived.

*Evaluation of Communication Changes*

Changes in communication score were expressed as differential percentage (ΔLRCS%):

$$\triangle LRCS\%=\frac{{LRCS}_{after}-{LRCS}_{before}}{{LRCS}_{before}}\times100\%$$

**Murine hypobaric-hypoxia exposure**

Male BALB/c mice aged 6–8 weeks were purchased from Chongqing Ensiweier Biotechnology Co., Ltd. and maintained under specific pathogen-free conditions. Thirty-two mice were randomly assigned to LB, LA, HB, and HA groups (n=8 per group).

HB and HA mice were continuously housed for 44 days in a hypobaric chamber (AVIC Fenglei Ordnance Co., Ltd., China) simulating an altitude of 5,800 m at a barometric pressure of approximately 50 kPa. Chamber pressure was gradually reduced from ambient pressure to the target level. Temperature was maintained at 22±2°C and relative humidity at 50±10%, with continuous airflow.

Following hypoxic exposure, LA and HA mice completed an exhaustive treadmill protocol at a 10° incline. After a 5-min warm-up at 10 m/min, running speed was increased by 1.5 m/min every minute until a maximum speed of 40 m/min was reached. Exhaustion was defined as remaining on the shock grid for >10 consecutive seconds without attempting to resume running.

**Murine sample preparation and flow cytometry**

Immediately after exhaustive exercise or at the corresponding resting time point, mice were anesthetized and peripheral blood was collected by retro-orbital bleeding. Spleens were harvested, mechanically dissociated, and passed through a 70-μm cell strainer. Erythrocytes were removed using RBC Lysis Buffer (MCE, HY-K3010).

Dead cells were excluded using Fixable Viability Dye 777 (BD Biosciences). Peripheral blood neutrophils were stained with CD11b-FITC (BD Biosciences, 557396), Ly6G-BV605 (563005), and CD62L-BV650 (564108).

Splenic B-cell analysis included CD19-RY610 (759096), CD138-BV421 (562610), GL7-AF647 (561529), and CD272/BTLA-RY586 (755174).

T-cell analysis included CD3-FITC (553061), CD4-BV480 (746631), CD8a-R718 (566985), CD44-RY775 (770596), CD62L-BV650 (564108), CD25-BV421 (564370), and PD-1-RB705 (758160).

Monocyte and dendritic-cell analysis included Ly6G-BV605 (563005), CD11b-FITC (557396), Ly6C-RY610 (759407), F4/80-BV421 (565411), CD11c-RY775 (771049), CD80-APC (560016), and CD86-BV650 (564200).

Flow-cytometric data were acquired using a FongCyte S four-laser flow cytometer (SinoCyte) and analyzed using FlowJo. Gating strategies are shown in the corresponding Supplementary Figures.

**Skeletal-muscle RNA extraction and library preparation**

Gastrocnemius muscles were rapidly dissected from HB and HA mice, snap-frozen in liquid nitrogen, and stored at −80°C. Total RNA was extracted using TRIzol reagent (Sangon Biotech, B511311) according to the manufacturer's instructions. RNA concentration was determined using the Qubit RNA Assay Kit on a Qubit 2.0 Fluorometer, and RNA integrity and genomic DNA contamination were assessed before library preparation.

Five biological replicates were analyzed per group (HB, n=5; HA, n=5). Poly(A)-containing mRNA was enriched using oligo(dT) magnetic beads and fragmented before first- and second-strand cDNA synthesis. Double-stranded cDNA underwent end repair, dA-tailing, adapter ligation, size selection, PCR amplification, and final purification. Qualified libraries were sequenced on an Illumina platform in paired-end 150-bp mode.

**Skeletal-muscle RNA-seq processing**

Raw sequencing reads were assessed using FastQC (v0.12.1). Adapter sequences and low-quality bases were removed using Trim Galore (v0.6.10, powered by Cutadapt) with a Phred quality threshold of 25, stringency of 15, and minimum retained read length of 120 bp.

Clean paired-end reads were aligned to the mouse reference genome GRCm38/mm10 using HISAT2 (v2.2.0). Alignment files were converted, sorted, and indexed using SAMtools (v1.6), and gene-level counts were generated using featureCounts (v2.0.6) against the GENCODE vM25 annotation.

Differential expression between HA and HB was analyzed using DESeq2 (v1.42.0). Genes with P<0.05 and |log2FC|>0.5 were considered differentially expressed. Upregulated and downregulated genes were analyzed separately for Gene Ontology enrichment using clusterProfiler (v4.10.0).

Immune-cell-associated signatures were estimated by ssGSEA using the GSVA package (v1.50.5). FPKM values were used as input, and immune-cell marker gene sets were obtained from the previously published compendium [50]. Differences between HB and HA were assessed using the Wilcoxon rank-sum test.

Supplementary Figures

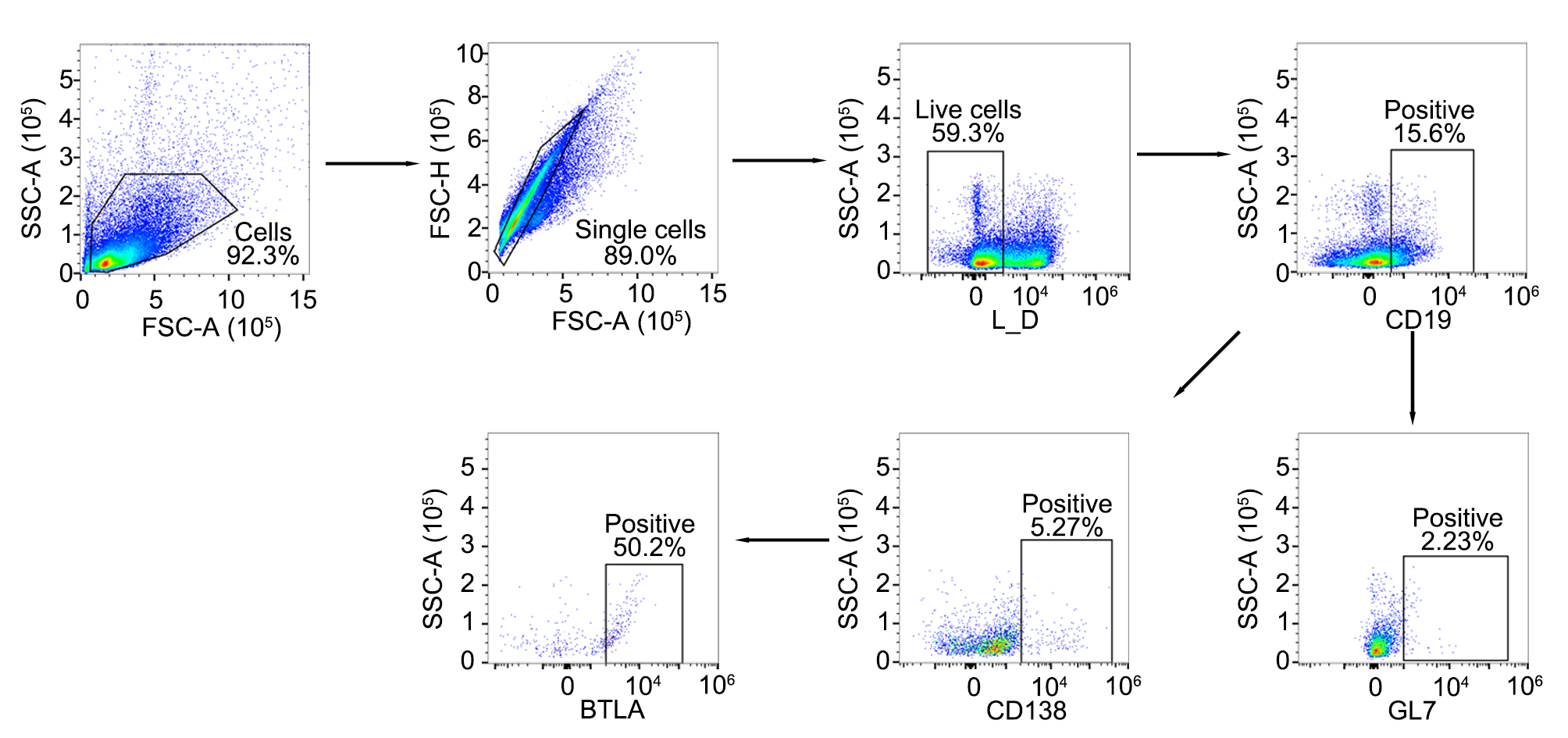

**Figure S1. Flow cytometry gating strategy for the identification of murine splenic B cell subsets and their functional phenotypes.**
Representative flow cytometry plots illustrating the sequential gating strategy. Debris was first excluded based on forward scatter (FSC-A) and side scatter (SSC-A) to identify total cells. Doublets were subsequently excluded using FSC-A versus FSC-H to select single cells. Dead cells were gated out using a Live/Dead (L_D) viability dye. Within the live single-cell population, total B cells were identified as CD19⁺. From the CD19⁺ gate, activated B cells and plasmablasts were identified based on the expression of GL7 and CD138, respectively. Finally, the surface expression of the inhibitory receptor BTLA was specifically evaluated within the gated CD138⁺ plasmablast population.

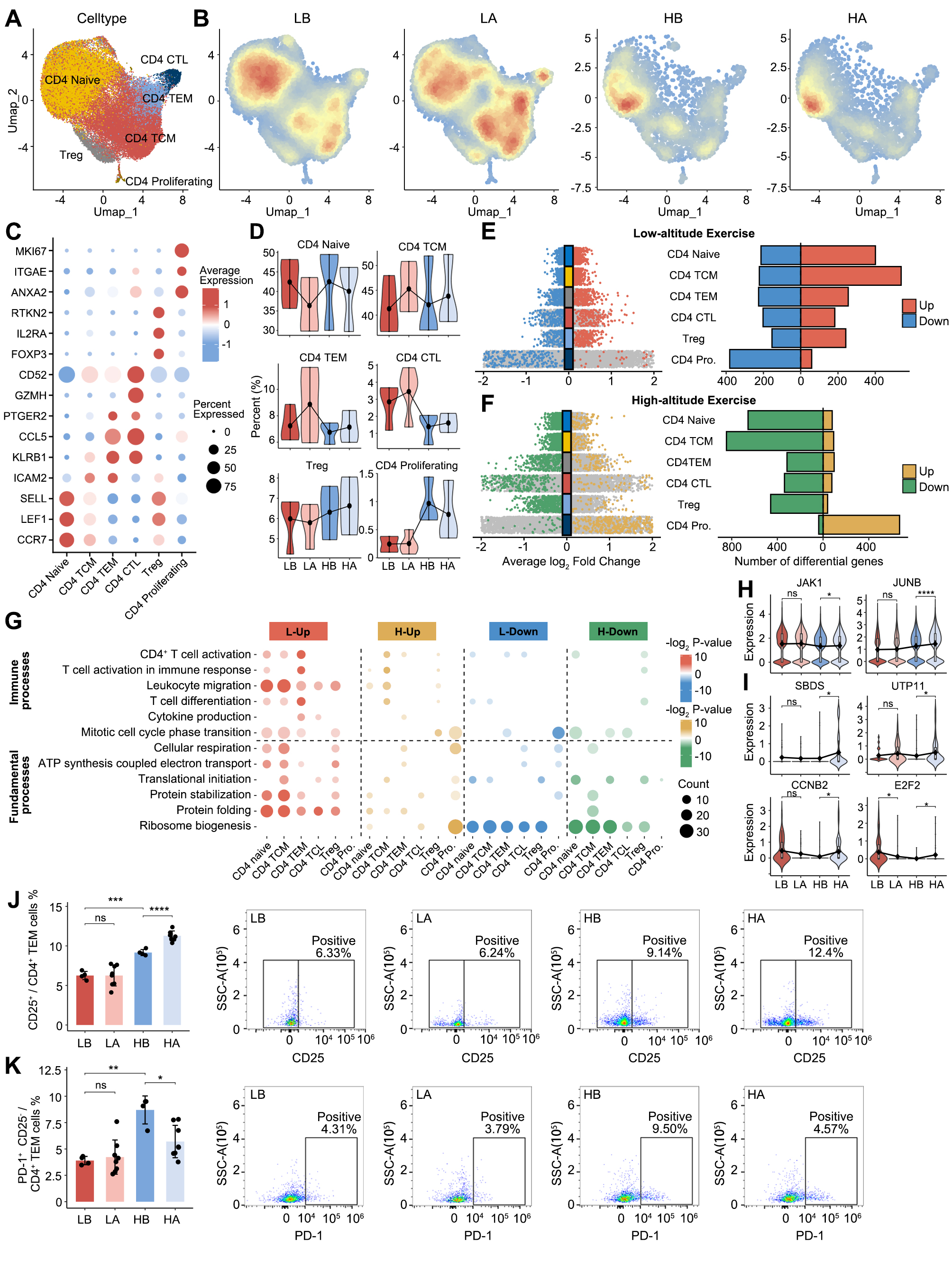

**Figure S2. Cellular and functional heterogeneity in the CD4^+^ T cells in response to exercise at low-altitude and high-altitude**

**A.** UMAP plot of the CD4⁺ T cell subtypes.
**B.** Cell density plots of CD4⁺ T cells across four experimental conditions (LB, LA, HB, HA), with color intensity proportional to local cell density.
**C.** Dot plots depicting the percentages and average expression levels of canonical marker genes in CD4⁺ T cell subtypes.
**D.** Violin plots showing the changes in the relative proportions of CD4⁺ T cell subtypes across four experimental conditions. Black dots and connecting lines represent the mean values.
**E.** Multi-group volcano plots displaying exercise-induced DEGs across CD4⁺ T cell subtypes at low altitude, with significantly upregulated (red) and downregulated (blue) genes indicated (left). Bar plot depicting the number of DEGs in each CD4⁺ T cell subtype (right).
**F.** Multi-group volcano plots displaying exercise-induced DEGs across CD4⁺ T cell subtypes at high altitude, with significantly upregulated (orange) and downregulated (green) genes indicated (left). Bar plot depicting the number of DEGs in each CD4⁺ T cell subtype (right).
**G.** Functional enrichment analysis of DEGs across CD4⁺ T cell subtypes under low-altitude and high-altitude exercise conditions. The bubble plot displays significantly enriched GO biological processes for four categories: L-Up (red), H-Up (orange), L-Down (blue), and H-Down (green). Color intensity represents -log2(*P*-value), and bubble size indicates the number of genes enriched in each GO term.
**H.** Violin plots showing expression levels of representative genes associated with T-cell activation (*JAK1*, *JUNB*) in CD4⁺ TCM cells across four experimental conditions (LB, LA, HB, HA).
**I.** Violin plots showing expression levels of representative genes involved in cell cycle regulation (*CCNB2*, *E2F2*) and ribosome biogenesis (*SBDS*, *UTP11*) in CD4⁺ proliferating cells across four experimental conditions.
**J.** Flow cytometry quantification (left) and representative dot plots (right) showing the percentage of activated CD25⁺ cells within the splenic CD4⁺ effector memory T (TEM) cell population across four experimental conditions in the murine model.
**K.** Flow cytometry quantification (left) and representative dot plots (right) showing the percentage of exhausted PD-1⁺ cells specifically within the resting (CD25⁻) CD4⁺ TEM cell population.

For H-K, statistical significance is indicated as follows: *p < 0.05, **p < 0.01, ***p < 0.001, ****p < 0.0001, ns = not significant. Bar graphs in J and K are presented as mean ± SD.

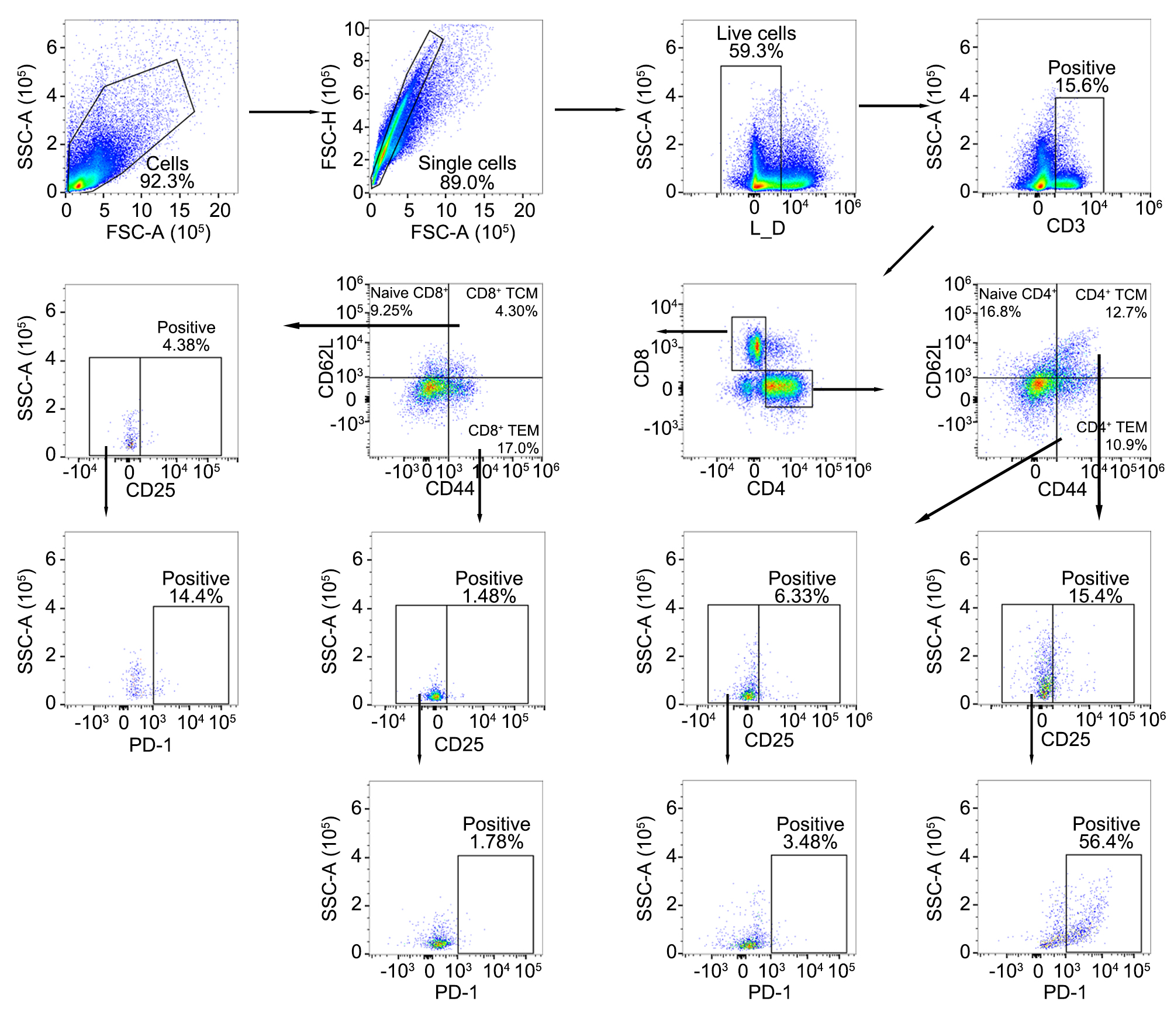

**Figure S3. Flow cytometry gating strategy for the identification of murine splenic CD4⁺ and CD8⁺ T cell subsets and their functional phenotypes.**
Representative flow cytometry plots illustrating the sequential gating strategy. Debris was first excluded based on forward scatter (FSC-A) and side scatter (SSC-A), followed by doublet exclusion using FSC-A versus FSC-H to select single cells. Dead cells were gated out using a Live/Dead (L_D) viability dye. Within the live single-cell population, total T cells were identified as CD3⁺. The CD3⁺ T cells were subsequently subdivided into CD4⁺ and CD8⁺ T cell lineages. Within both the CD4⁺ and CD8⁺ compartments, T cell subsets were further defined based on the differential expression of CD44 and CD62L: naive T cells (CD62L⁺ CD44⁻), central memory T cells (TCM; CD62L⁺ CD44⁺), and effector memory T cells (TEM; CD62L⁻ CD44⁺). Finally, the surface expression of the activation marker CD25 and the inhibitory receptor PD-1 was evaluated downstream within the specific memory T cell compartments (TCM and TEM) for both CD4⁺ and CD8⁺ lineages.

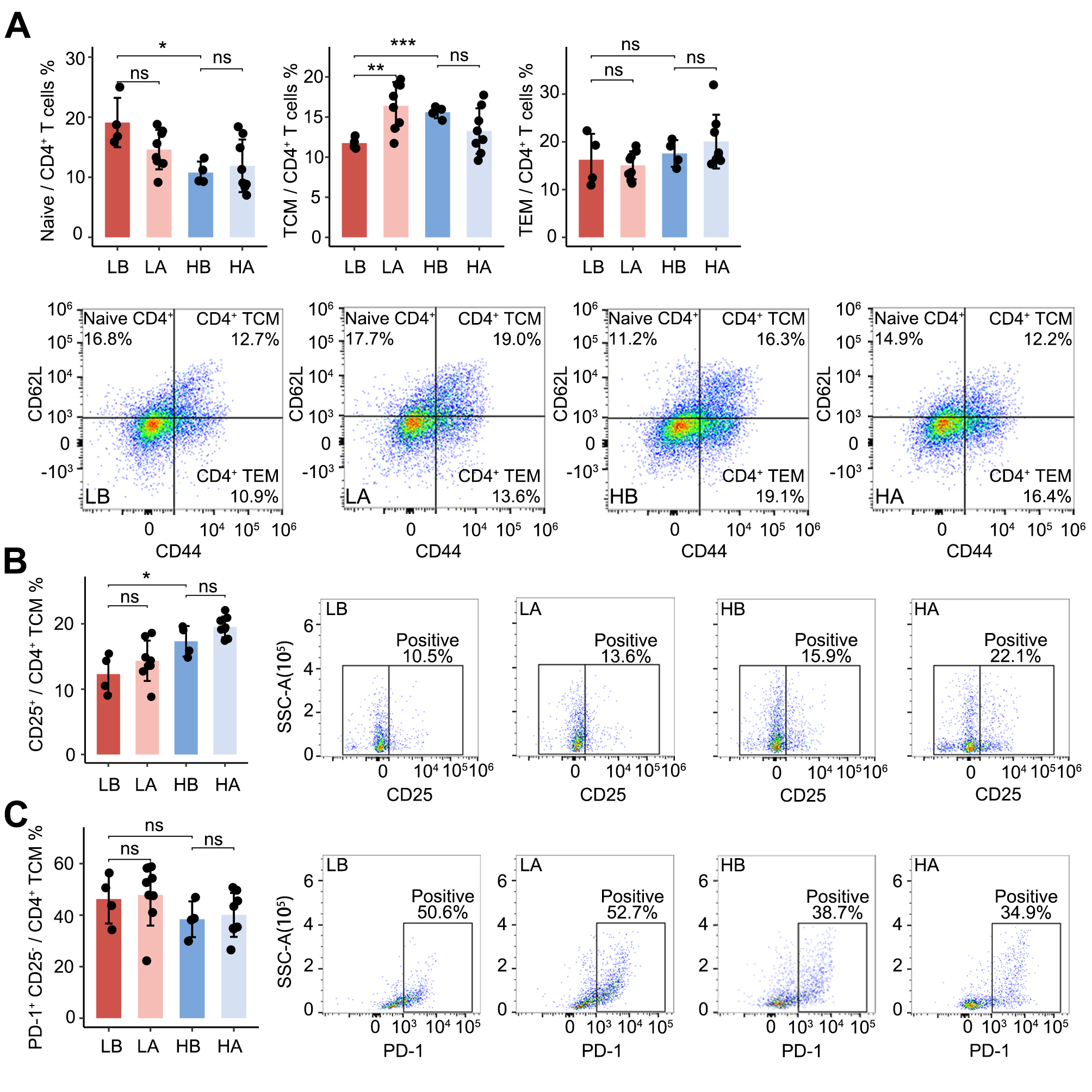

**Figure S4. Phenotypic and functional characterization of splenic CD4⁺ T cell subsets across experimental conditions in the murine model.**
**A.** Flow cytometry quantification (top) and representative pseudocolor plots (bottom) showing the relative proportions of Naive (CD62L⁺ CD44⁻), central memory (TCM; CD62L⁺ CD44⁺), and effector memory (TEM; CD62L⁻ CD44⁺) subsets within the total splenic CD4⁺ T cell population across four experimental conditions (LB, LA, HB, HA).
**B.** Flow cytometry quantification (left) and representative dot plots (right) showing the percentage of activated CD25⁺ cells specifically within the CD4⁺ TCM population.
**C.** Flow cytometry quantification (left) and representative dot plots (right) showing the percentage of exhausted PD-1⁺ cells evaluated within the resting (CD25⁻) fraction of the CD4⁺ TCM population.

Statistical significance is indicated as follows: *p < 0.05, **p < 0.01, ***p < 0.001, ns = not significant. All bar graphs are presented as mean ± SD.

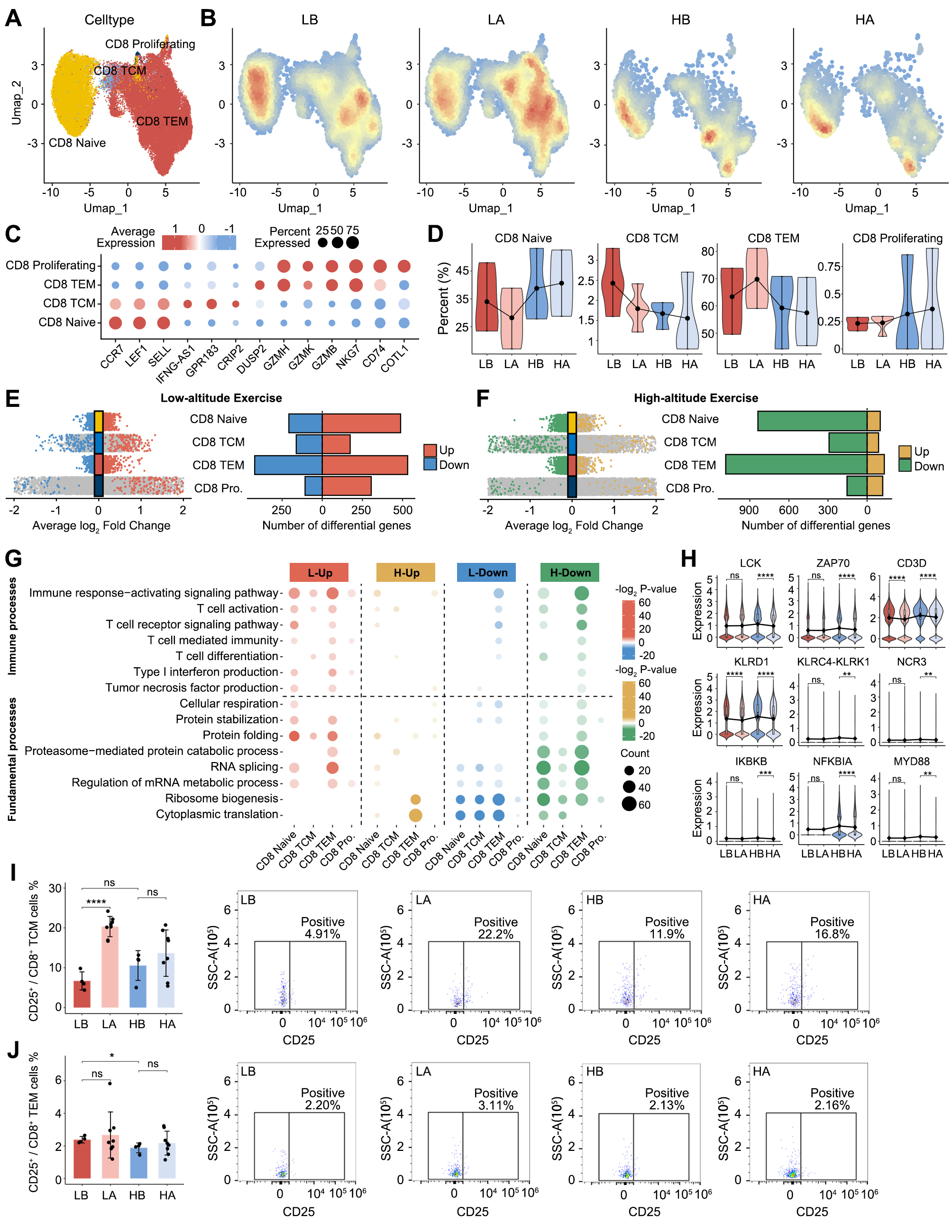

**Figure S5. Cellular and functional heterogeneity in the CD8^+^ T cell in response to exercise at low-altitude and high-altitude**

**A.** UMAP plot of the CD8^+^ T cell subtypes.

**B.** Cell density plots of CD8^+^ T cells across four experimental conditions (LB, LA, HB, HA), with color intensity proportional to local cell density.

**C.** Dot plots depicting the percentages and average expressions of marker genes in CD8^+^ T cell subtypes.

**D.** Violin plots showing the changes in the relative proportions of CD8^+^ T cell subtypes across four experimental conditions (LB, LA, HB, HA).

**E.** Multi-group volcano plots displaying exercise-induced DEGs across CD8^+^ T cell subtypes at low-altitude, with significantly upregulated (red) and downregulated (blue) genes indicated (left). Bar plot depicting the number of DEGs in each CD8^+^ T cell subtype (right).

**F.** Multi-group volcano plots displaying exercise-induced DEGs across CD8^+^ T cell subtypes at high-altitude, with significantly upregulated (orange) and downregulated (green) genes indicated. Bar plot depicting the number of DEGs in each CD8^+^ T cell subtype (right).

**G.** Functional enrichment analysis of DEG across CD8^+^ T cell subtypes under low-altitude and high-altitude exercise conditions. The bubble plot displays significantly enriched GO biological processes for four categories: L-Up (red), H-Up (orange), L-Down (blue), and H-Down (green). Color intensity represents -log_2_(P-value), and bubble size indicates the number of genes enriched in each GO term.

**H.** Violin plots showing representative genes involved in TCR signaling (LCK, ZAP70, CD3D), cytotoxicity (KLRD1, KLRC4–KLRK1, NCR3), and NF-κB pathways (IKBKB, NFKBIA, MYD88) in CD8⁺ TEM cells across four experimental conditions (LB, LA, HB, HA).

**I.** Flow cytometry quantification (left) and representative dot plots (right) showing the percentage of activated CD25⁺ cells within the splenic CD8⁺ central memory T (TCM) cell population across four experimental conditions in the murine model.
**J.** Flow cytometry quantification (left) and representative dot plots (right) showing the percentage of activated CD25⁺ cells within the splenic CD8⁺ effector memory T (TEM) cell population.

For H-J, statistical significance is indicated as follows: *p < 0.05, **p < 0.01, ***p < 0.001, ****p < 0.0001, ns = not significant. Bar graphs in I and J are presented as mean ± SD.

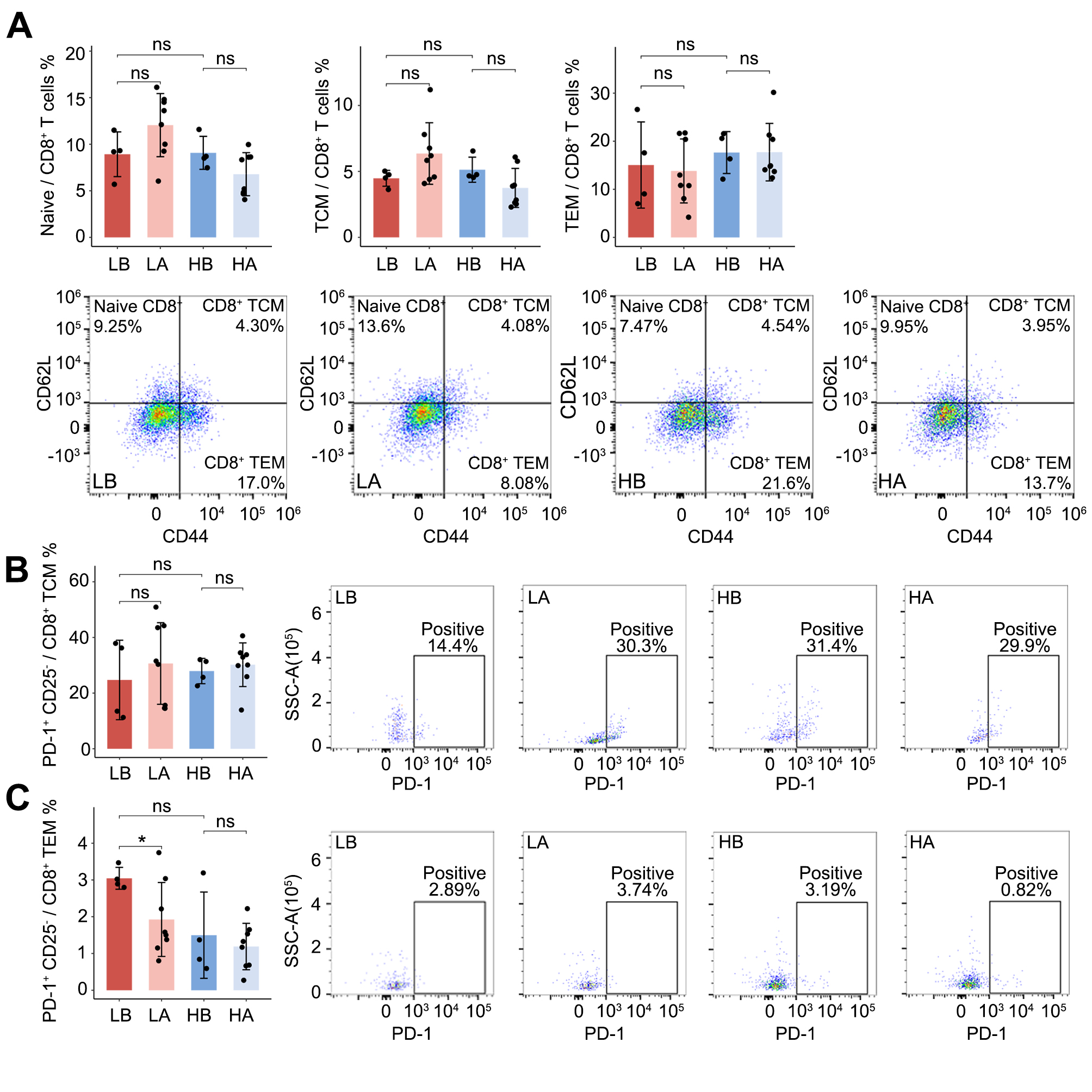

**Figure S6. Phenotypic and functional characterization of splenic CD8⁺ T cell subsets across experimental conditions in the murine model.**
**A.** Flow cytometry quantification (top) and representative pseudocolor plots (bottom) showing the relative proportions of Naive (CD62L⁺ CD44⁻), central memory (TCM; CD62L⁺ CD44⁺), and effector memory (TEM; CD62L⁻ CD44⁺) subsets within the total splenic CD8⁺ T cell population across four experimental conditions (LB, LA, HB, HA).
**B.** Flow cytometry quantification (left) and representative dot plots (right) showing the percentage of exhausted PD-1⁺ cells evaluated specifically within the resting (CD25⁻) fraction of the CD8⁺ TCM population.
**C.** Flow cytometry quantification (left) and representative dot plots (right) showing the percentage of exhausted PD-1⁺ cells evaluated specifically within the resting (CD25⁻) fraction of the CD8⁺ TEM population.

Statistical significance is indicated as follows: *P < 0.05, ns = not significant. All bar graphs are presented as mean ± SD.

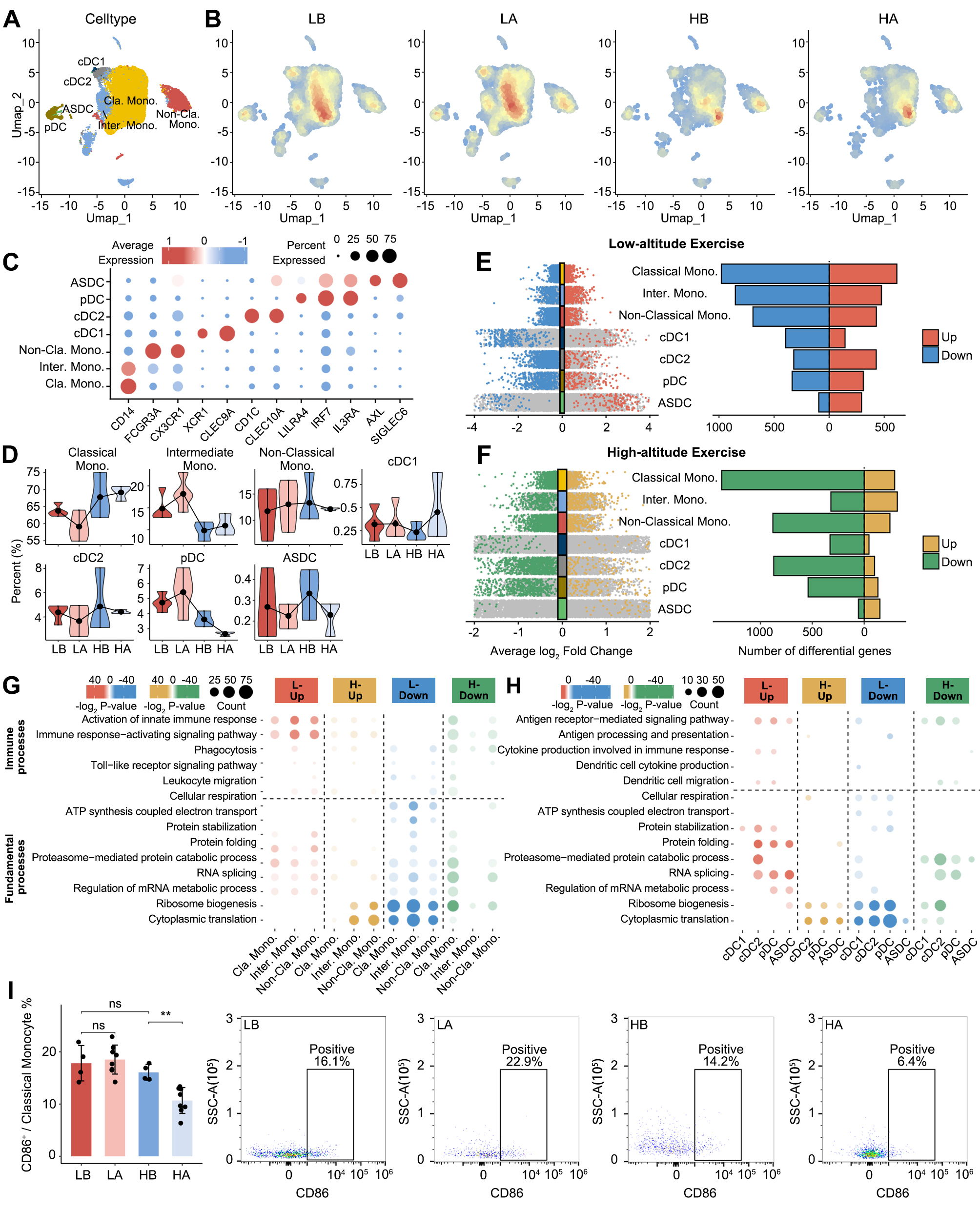

**Figure S7. Cellular and functional heterogeneity in the monocyte and DCs in response to exercise at low-altitude and high-altitude**

A. UMAP plot of the monocyte and DCs subtypes.

B. Cell density plots of monocyte and DCs across four experimental conditions (LB, LA, HB, HA), with color intensity proportional to local cell density.

**C.** Dot plots depicting the percentages and average expressions of marker genes in monocyte and DCs subtypes.

**D.** Violin plots showing the changes in the relative proportions of monocyte and DCs subtypes across four experimental conditions (LB, LA, HB, HA).

**E.** Multi-group volcano plots displaying exercise-induced DEGs across monocyte and DCs subtypes at low-altitude, with significantly upregulated (red) and downregulated (blue) genes indicated (left). Bar plot depicting the number of DEGs in each monocyte and DCs subtype (right).

**F.** Multi-group volcano plots displaying exercise-induced DEGs across monocyte and DCs subtypes at high-altitude, with significantly upregulated (orange) and downregulated (green) genes indicated. Bar plot depicting the number of DEGs in each monocyte and DCs subtype (right).

**G-H.** Functional enrichment analysis of DEG across monocyte subtypes (G) and DCs subtypes (H) under low-altitude and high-altitude exercise conditions. The bubble plot displays significantly enriched GO biological processes for four categories: L-Up (red), H-Up (orange), L-Down (blue), and H-Down (green). Color intensity represents -log_2_(P-value), and bubble size indicates the number of genes enriched in each GO term.

**I.** Flow cytometry quantification (left) and representative dot plots (right) showing the percentage of CD86⁺ cells within the classical monocyte population across four experimental conditions in the murine model.

For I, statistical significance is indicated as follows: *p < 0.01, ns = not significant. The bar graph is presented as mean ± SD.

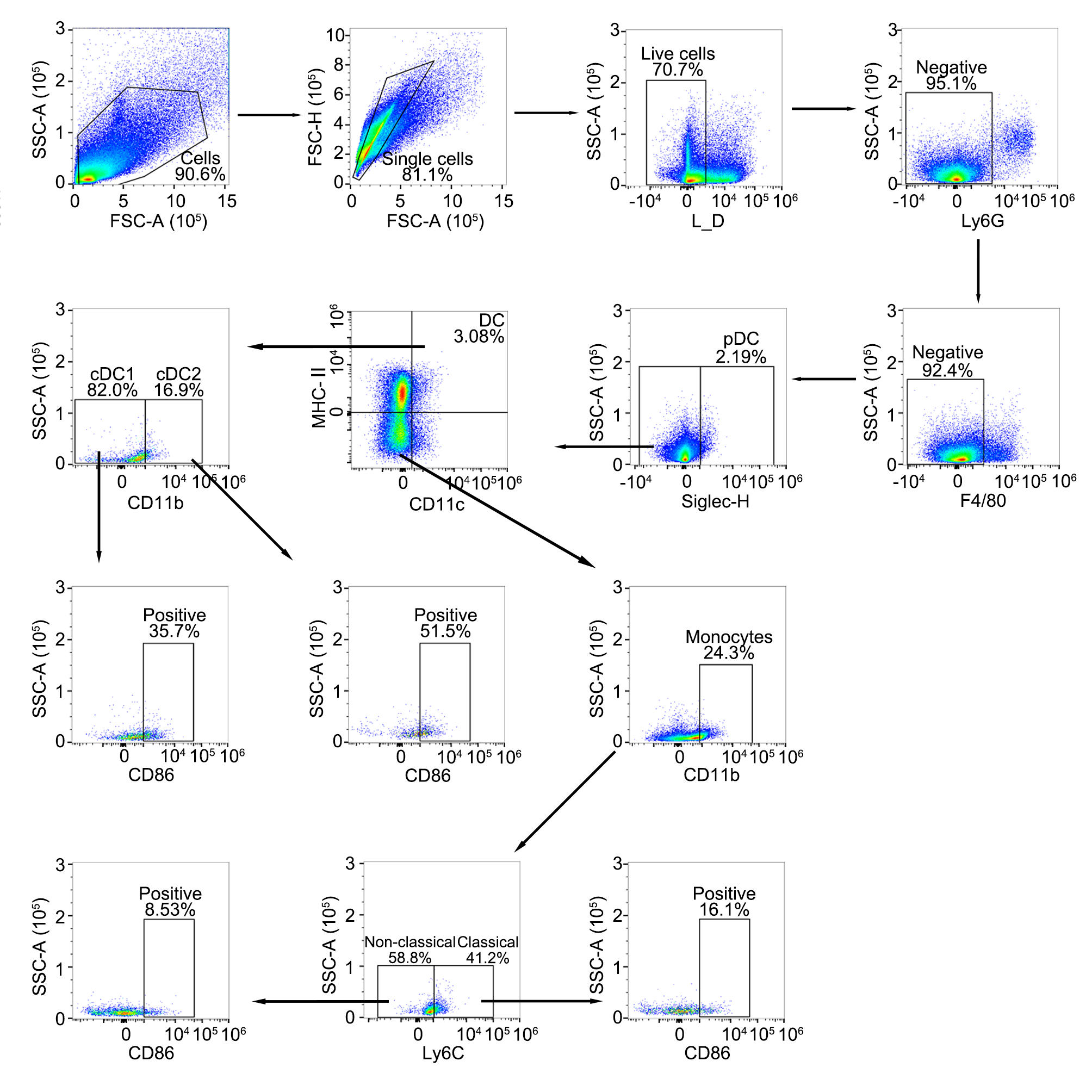

**Figure S8. Flow cytometry gating strategy for the identification of murine monocyte and dendritic cell (DC) subsets and their activation status.**
Representative flow cytometry plots illustrating the sequential gating strategy. Debris and doublets were excluded using forward scatter (FSC) and side scatter (SSC) parameters, followed by the exclusion of dead cells using a Live/Dead (L_D) viability dye. To specifically identify mononuclear phagocyte subsets, neutrophils and macrophages were sequentially gated out from the live single-cell population by selecting the Ly6G⁻ and F4/80⁻ fractions, respectively. Within this Ly6G⁻ F4/80⁻ population, plasmacytoid DCs (pDCs) were identified as Siglec-H⁺. Conventional DCs (cDCs) were identified as CD11c⁺ MHC-II⁺, which were further subdivided into cDC1 (CD11b⁻) and cDC2 (CD11b⁺) subsets. Total monocytes were identified from the non-cDC fraction as CD11b⁺ cells. These monocytes were subsequently classified into classical (Ly6C⁺) and non-classical (Ly6C⁻) subsets based on Ly6C expression. Finally, the surface expression of the co-stimulatory molecule CD86 was evaluated downstream within each specific DC (cDC1, cDC2) and monocyte (classical, non-classical) subset.

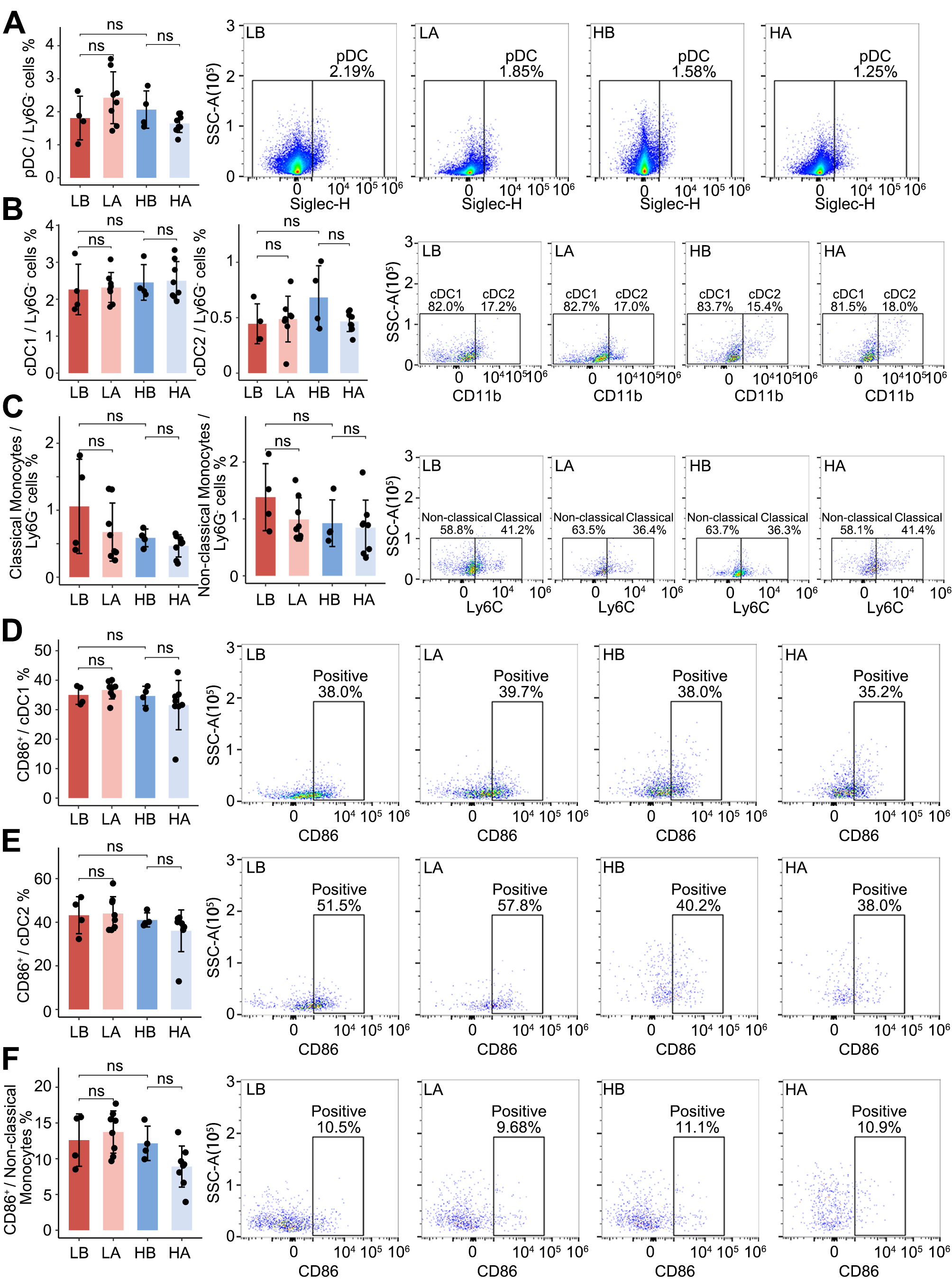

**Figure S9. Phenotypic and functional characterization of murine monocyte and dendritic cell (DC) subsets across experimental conditions.**
**A.** Flow cytometry quantification of pDCs as a percentage of total Ly6G⁻ cells (left), alongside representative pseudocolor plots showing the Siglec-H⁺ pDC gating (right).
**B.** Flow cytometry quantification of cDC1 and cDC2 subsets as a percentage of total Ly6G⁻ cells (left). Representative dot plots illustrate the relative distribution of cDC1 (CD11b⁻) and cDC2 (CD11b⁺) subsets within the pre-gated CD11c⁺ MHC-II⁺ cDC population (right).
**C.** Flow cytometry quantification of classical and non-classical monocytes as a percentage of total Ly6G⁻ cells (left). Representative dot plots illustrate the relative distribution of non-classical (Ly6C⁻) and classical (Ly6C⁺) subsets within the pre-gated total monocyte population (right).
**D.** Flow cytometry quantification (left) and representative dot plots (right) showing the percentage of CD86⁺ cells evaluated within the cDC1 population across four experimental conditions.
**E.** Flow cytometry quantification (left) and representative dot plots (right) showing the percentage of CD86⁺ cells evaluated within the cDC2 population.
**F.** Flow cytometry quantification (left) and representative dot plots (right) showing the percentage of CD86⁺ cells evaluated within the non-classical monocyte population.

Statistical significance is indicated as follows: ns = not significant. All bar graphs are presented as mean ± SD.

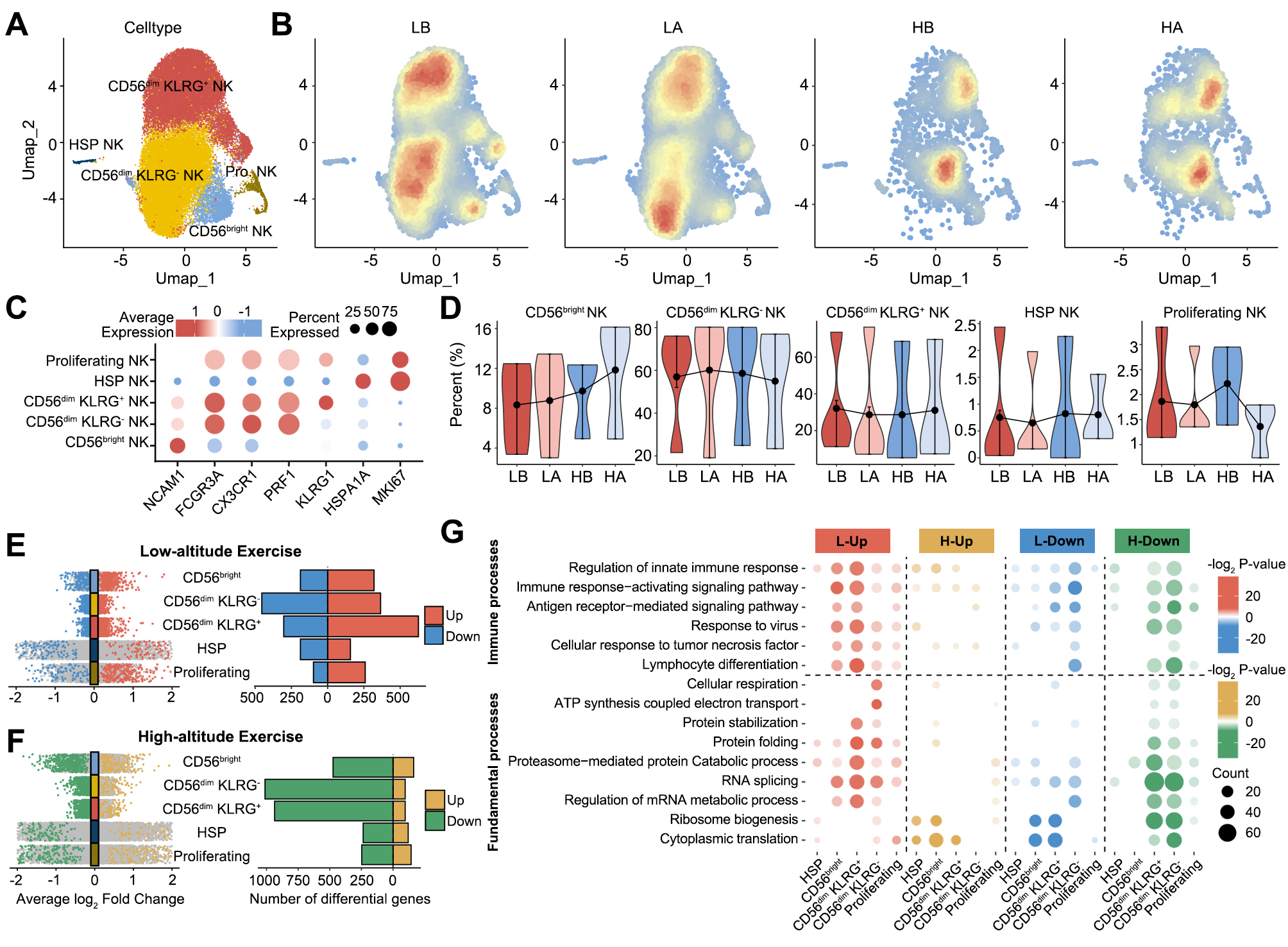

**Figure S10. Cellular and functional heterogeneity in the NK cell in response to exercise at low-altitude and high-altitude**

**A.** UMAP plot of the NK cell subtypes.

**B.** Cell density plots of NK cells across four experimental conditions (LB, LA, HB, HA), with color intensity proportional to local cell density.

**C.** Dot plots depicting the percentages and average expressions of marker genes in NK cell subtypes.

**D.** Violin plots showing the changes in the relative proportions of NK cell subtypes across four experimental conditions (LB, LA, HB, HA).

**E.** Multi-group volcano plots displaying exercise-induced DEGs across NK cell subtypes at low-altitude, with significantly upregulated (red) and downregulated (blue) genes indicated (left). Bar plot depicting the number of DEGs in each NK cell subtype (right).

**F.** Multi-group volcano plots displaying exercise-induced DEGs across NK cell subtypes at high-altitude, with significantly upregulated (orange) and downregulated (green) genes indicated. Bar plot depicting the number of DEGs in each NK cell subtype (right).

**G.** Functional enrichment analysis of DEGs across NK cell subtypes under low-altitude and high-altitude exercise conditions. The bubble plot displays significantly enriched GO biological processes for four categories: L-Up (red), H-Up (orange), L-Down (blue), and H-Down (green). Color intensity represents -log_2_(P-value), and bubble size indicates the number of genes enriched in each GO term.

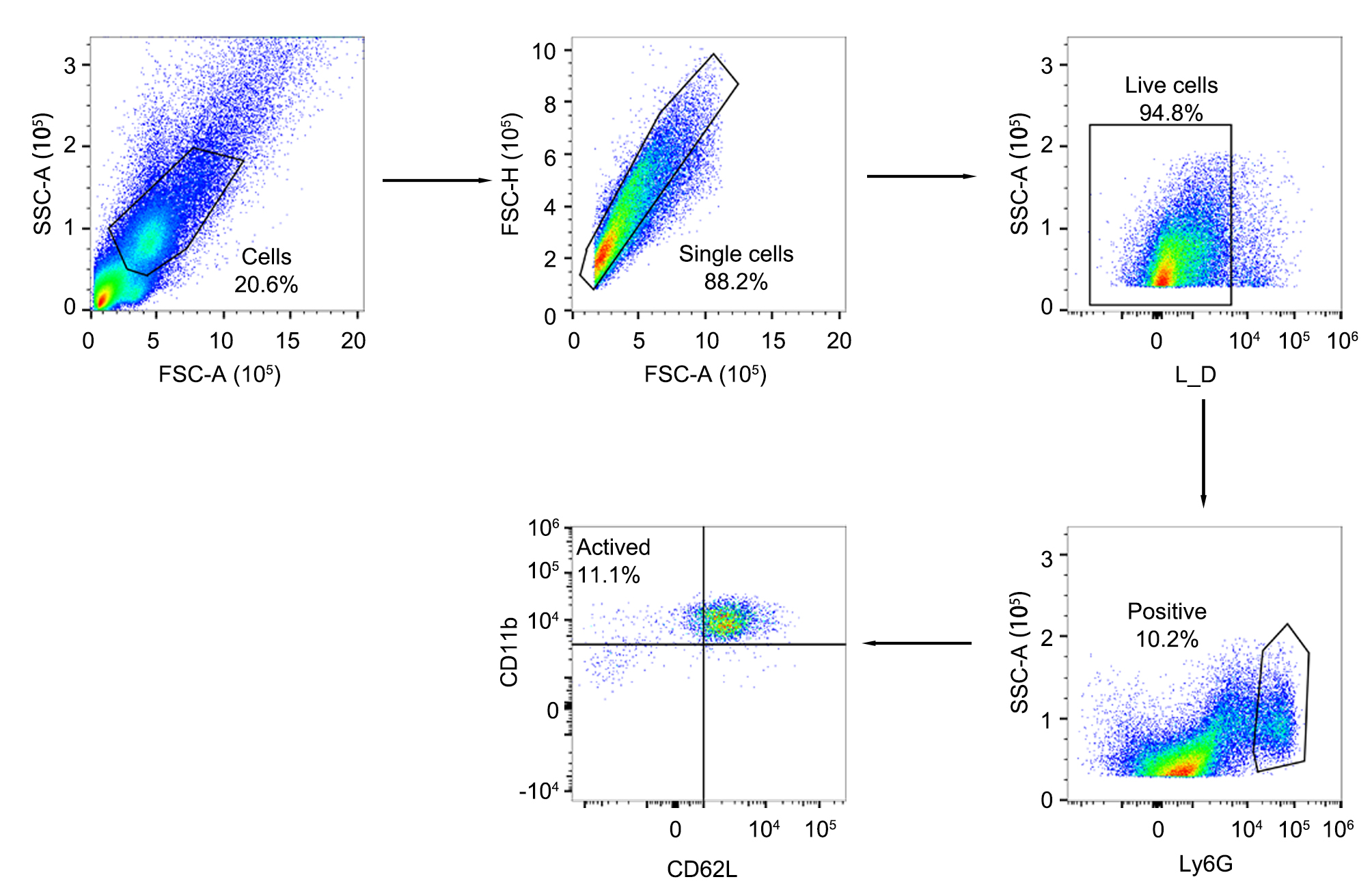

**Figure S11. Flow cytometry gating strategy for the identification of murine activated mature neutrophils.**
Representative flow cytometry plots illustrating the sequential gating strategy. Debris and doublets were sequentially excluded using forward scatter (FSC) and side scatter (SSC) parameters (FSC-A vs. SSC-A and FSC-A vs. FSC-H, respectively). Dead cells were then gated out using a Live/Dead (L_D) viability dye. Within the live single-cell population, total neutrophils were identified based on the positive expression of Ly6G. Finally, the activated mature neutrophil subset was defined within the Ly6G⁺ parent population based on the shedding of CD62L and the high expression of CD11b (identified as the CD62L⁻ CD11b⁺ fraction in the upper left quadrant).

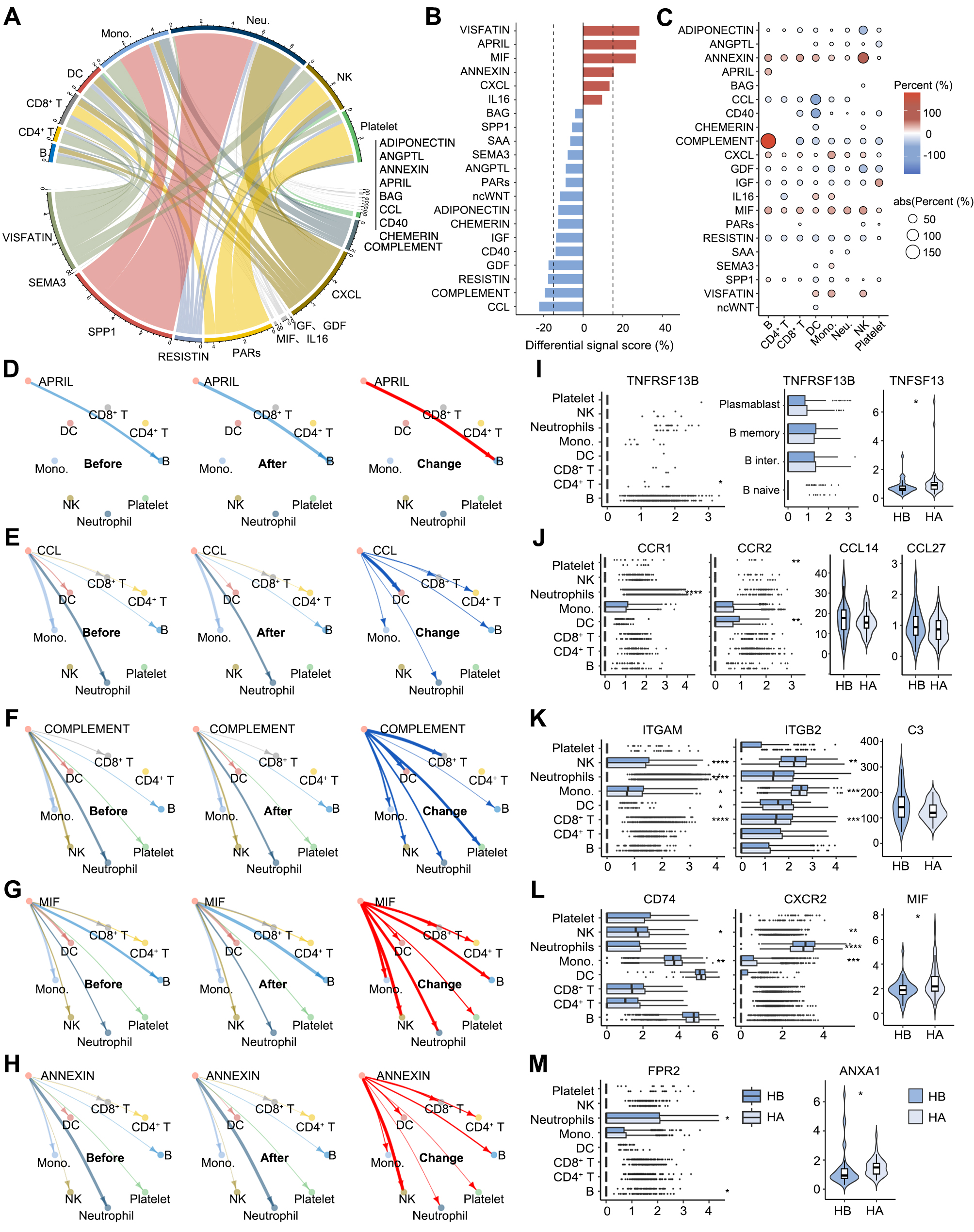

**Figure S12. Integrated transcriptomic-proteomic analysis reveals remodeling of communication networks.**

**A.** Chord diagram depicting the global communication landscape between 21 signaling pathways (left hemisphere) and eight major immune cell types (right hemisphere).

**B.** Bar plot showing the percentage change in the total communication strength of signaling pathways following high-altitude exercise.

**C.** Dot plot displaying the change in pathway-level communication strength for each immune cell type. Color represents the percentage change, and dot size represents the absolute value of the change.

**D-H.** Circle plots visualizing the communication networks before (HB) and after (HA) high-altitude exercise, alongside the differential changes (Change) for key signaling pathways: (D) APRIL, (E) CCL, (F) COMPLEMENT, (G) MIF, and (H) ANNEXIN. Red arrows indicate upregulated signaling, and blue arrows indicate downregulated signaling.

**I-M.** Corresponding molecular alterations for each pathway. Box plots show the scRNA-seq expression levels of specific receptors across immune subsets, and violin plots show the plasma protein abundance of the corresponding circulating ligands before (HB) and after (HA) high-altitude exercise. Panels correspond to the (I) APRIL pathway (TNFRSF13B; TNFSF13), (J) CCL pathway (CCR1, CCR2; CCL14, CCL27), (K) COMPLEMENT pathway (ITGAM, ITGB2; C3), (L) MIF pathway (CD74, CXCR2; MIF), and (M) ANNEXIN pathway (FPR2; ANXA1).

For I-M, statistical significance is indicated as follows: *p < 0.05, **p < 0.01, ***p < 0.001, ****p < 0.0001.
